# Polygenic risk scores and Mendelian randomization reveal circadian genetic contributions to idiopathic hypersomnia

**DOI:** 10.64898/2026.01.06.26343504

**Authors:** Taku Miyagawa, Mihoko Shimada, Kotomi Tanida, Nozomu Kotorii, Takao Kato, Tatayu Kotorii, Yu Ariyoshi, Hiroshi Hiejima, Motohiro Ozone, Naohisa Uchimura, Azusa Ikegami, Kazuhiko Kume, Takashi Kanbayashi, Aya Imanishi, Yuichi Kamei, Akiko Hida, Yamato Wada, Kenji Kuroda, Masayuki Miyamoto, Koichi Hirata, Masanori Takami, Naoto Yamada, Masako Okawa, Naoto Omata, Hideaki Kondo, Tohru Kodama, Yuichi Inoue, Kazuo Mishima, Katsushi Tokunaga, Makoto Honda

## Abstract

Idiopathic hypersomnia (IH) is a rare and heterogeneous sleep disorder characterized by excessive daytime sleepiness. We aimed to stratify IH patients based on polygenic risk scores (PRSs) for sleep-related traits and explored the underlying genetic predispositions. Genome-wide single-nucleotide polymorphism data from 303 Japanese IH patients and 2,918 controls were analyzed. PRSs were calculated for chronotype (morningness/eveningness), daytime napping, sleep duration, and insomnia using publicly available base data. Patients in the extreme PRS tails were examined for clinical phenotypes. Causal inference was evaluated via Mendelian randomization (MR). IH patients were significantly enriched in the top PRS percentiles for eveningness (top 0.5%: odds ratio [OR] = 3.40, P = 3.9×10^−5^; top 1%: OR = 2.48, P = 1.9×10^−4^) and daytime napping (top 0.5%: OR = 3.80; P = 1.6×10^−5^), with substantially elevated ORs. Patients with high morningness PRS exhibited increased slow-wave sleep near wake time, frequently observed in patients with IH. MR analysis supported a causal relationship between eveningness and IH (inverse-variance weighted, UK Biobank: P = 1.3×10^−3^; replicated in 23andMe P = 0.037). This causal association was consistently observed even among a subgroup of IH patients who showed ≥660 minutes of total sleep time in 24-hour polysomnography. PRS analysis identified significant associations between IH and sleep traits such as eveningness and daytime napping, with notably large effect sizes. Importantly, MR analysis further suggested a causal role for eveningness in IH pathogenesis. These findings highlight distinct genetic subtypes within IH and reinforce the relevance of circadian misalignment in its pathophysiology.

## Introduction

Idiopathic hypersomnia (IH) is a rare heterogeneous sleep disorder characterized by excessive daytime sleepiness with long and unrefreshing naps, prolonged nocturnal sleep, and sleep drunkenness(*1–4*). Symptoms generally begin and develop in adolescence or early adulthood. IH patients often have a family history of similar conditions(*5–7*), suggesting genetic involvement in IH. Recent genome-wide association studies (GWASs) and human leukocyte antigen (HLA) analyses identified a large number of genetic variants associated with narcolepsy type1(*8, 9*), whereas to date, only one rare variant in the *prepro-orexin* gene has been reported to be associated with IH(*10*). The rarity and heterogeneity of IH pose significant challenges in identifying associated genetic variants because the causes of IH can vary significantly between patients.

In recent years, the application of polygenic risk score (PRS) analysis for the prediction of disease onset has increased. Since its application for cardiovascular disease research, recognition of the effectiveness of PRS analysis has increased. Based on the results of a GWAS of over 200,000 individuals for risk of cardiovascular disease (base set), PRSs of individuals in a separate cohort (target set) were calculated, and individuals with the top 1% of PRSs had approximately a 5-fold increased risk of cardiovascular disease(*11, 12*). This groundbreaking finding quickly focused attention on PRS analysis. We attempted to classify subtypes of the heterogeneous disorder IH using an analysis method based on PRS. In conventional PRS analysis for estimating disease risk at the individual level, base data and target data are derived from genome-wide SNP data for the same disease. The aim of our study, therefore, was to identify disease subtypes by implementing a PRS analysis with a concept, as described below. The PGS Catalog provides base data calculated from data derived from various biobanks and disease databanks(*13, 14*). Among them are base data derived from sleep phenotypes and SNP data of hundreds of thousands of individuals, such as those from the UK Biobank. We hypothesized that these base data pertaining to sleep phenotypes could be used to stratify the target data (our IH patients) based on their PRSs for the sleep phenotypes. As IH is a rare sleep disorder, performing a large GWAS for IH would be challenging. Therefore, this PRS analysis utilizing base data from a large GWAS of sleep phenotypes is considered a practical and effective method for studying the genetic background of IH. In this study, we analyzed SNP data for IH patients and controls as the target set, identifying individuals in the top and bottom percentiles for each sleep phenotype. By examining the overrepresentation of IH patients in these top and bottom ranges compared with controls, we attempted to determine whether IH patients exhibit distinct genetic predispositions related to specific sleep traits. This approach enabled the stratification of IH patients from the perspective of genetic sleep traits. In addition, clinical characteristics of IH patients with extreme PRSs were assessed using both objective and subjective sleep parameters. Finally, a two-sample Mendelian randomization (MR) analysis was conducted to evaluate the potential causal relationship between the identified sleep trait and IH.

## Methods

### Subjects, Genotyping, and Imputation IH

The sample set included 306 patients with IH and 2918 healthy controls in the Japanese population. Physician sleep specialists diagnosed patients with IH according to the International Classification of Sleep Disorders, Third Edition, Text Revision (ICSD-3-TR)(*4*). Briefly, patients were diagnosed with IH if they met at least one of the following criteria: (1) a mean sleep latency of ≤8 min and ≤1 sleep-onset REM period (SOREMP) on the multiple sleep latency test (MSLT), or (2) a total sleep time of ≥660 min on 24-hour polysomnography (PSG). In addition, standard PSG and MSLT findings were inconsistent with the diagnostic criteria for narcolepsy type 1or type 2. According to the updated ICSD-3-TR, individuals with ≥660 minutes of total sleep time on 24-h PSG can be diagnosed with IH even if their MSLT shows a mean sleep latency >8 min irrespective of SOREMP occurrence, provided that findings do not meet criteria for narcolepsy type 1or type 2.

Genome-wide SNP typing was performed in 85 IH patients and 1374 healthy controls using the Genome-Wide Human SNP Array 6.0 and in 221 IH patients and 1544 healthy controls using the Axiom Genome-Wide ASI 1 Array Plate, according to the manufacturer’s protocols (Affymetrix Inc., Santa Clara, CA). The 85 IH patients and 1374 healthy controls genotyped using the Genome-Wide Human SNP Array 6.0 were obtained from a previous study(*15*). The 1544 healthy controls genotyped using the Axiom Genome-Wide ASI 1 Array Plate were obtained from the NBDC Human Database (accession numbers hum0082.v2 and hum0136.v3; https://humandbs.dbcls.jp/). Data generated using the Genome-Wide Human SNP Array 6.0 and Axiom Genome-Wide ASI 1 Array were analyzed using the GeneChip Operating Software and Genotyping Console 4.0 and the Axiom Analysis Suite, respectively (Affymetrix Inc.). We included SNPs that showed genotyping call rates >97%, minor allele frequencies (MAFs) >1%, and Hardy-Weinberg equilibrium (HWE) *P* values >10^−3^. Principal component analysis (PCA) was performed using PLINK 1.9 to evaluate population stratification among the study subjects genotyped using the abovementioned SNP arrays. The PCA examined 45 JPT (Japanese in Tokyo, Japan), 90 CEU (Utah residents with Northern and Western European ancestry from the CEPH collection), and 45 CHB (Han Chinese in Beijing, China) derived from the International HapMap project. One IH patient who was genotyped using the Axiom Genome-Wide ASI 1 Array Plate and located outside the East Asian cluster (JPT + CHB) was excluded. Unknown familial relationships among subjects were assessed using PIHAT values calculated using PLINK 1.9. Three IH patients who were genotyped using the Axiom Genome-Wide ASI 1 Array Plate and had PIHAT values >0.125 were excluded due to possible familial relationships. After quality control, genotype data from a total of 303 IH patients (85 genotyped using the Genome-Wide Human SNP Array 6.0 and 218 genotyped using the Axiom Genome-Wide ASI 1 Array Plate) and 2918 healthy controls (1374 genotyped using the Genome-Wide Human SNP Array 6.0 and 1544 genotyped using the Axiom Genome-Wide ASI 1 Array Plate) were retained for subsequent analyses. An imputation analysis was also performed to evaluate associations between ungenotyped genetic variants. Genotype phasing was performed using Eagle, and genotype imputation was carried out using Minimac4 with the 1000 Genomes Project Phase 3 multi-ethnic reference panel (GRCh37/hg19)(*16, 17*). Genetic variants with an imputation quality score (Rsq) >0.3, genotyping call rate >97%, MAF >1%, and HWE *P* value >10^−6^ were included. The imputed genotype data were used as the target dataset for PRS analysis and outcome data for MR analysis.

Of the 303 IH patients, 59 underwent 24-hour PSG, of whom 57 met the ICSD-3-TR criterion of total sleep time ≥660 minutes. The remaining two, along with 244 other patients (total n = 246), were diagnosed based on MSLT findings. Clinical data, including details regarding symptoms and conventional sleep parameters from standard PSG and MSLT were collected for IH patients for whom additional data were available. The sleep-corrected mid-sleep on free days (MSFsc) was used as an indicator for chronotype. The Japanese version of the Epworth Sleepiness Scale (JESS) was employed as a subjective measure of daytime sleepiness in the enrolled patients(*18*). This study was approved by the Research Ethics Committee of Tokyo Metropolitan Institute of Medical Science (21–10) and the Research Ethics Committee of the Institute of Neuropsychiatry (188–5). All subjects provided written informed consent. Clinical and demographic characteristics of the IH patients are summarized in Table 1 and Supplementary Tables S1 and S2.

**Table 1.** Demographic and sleep-related characteristics of patients with idiopathic hypersomnia (IH).

| Variable | N | Mean | SD or SE |
| --- | --- | --- | --- |
| Age, years | 303 | 25.71 | 8.95 |
| Sex, % female | 303 | 56.2% | 2.8%* |
| BMI | 272 | 21.25 | 3.15 |
| HLA-DQB1*06:02 positivity, % | 301 | 10.9% | 1.7%* |
| Sleep latency (MSLT), min | 246 | 6.07 | 4.12 |
| SOREMP, number | 246 | 0.28 | 0.53 |
| Sleep efficiency, % | 247 | 88.5% | 11.2% |
| Apnea hypopnea index, events/h | 250 | 2.36 | 2.84 |
| Sleep latency (PSG), min | 250 | 16.47 | 34.29 |
| PLMI, events/h | 248 | 0.89 | 2.03 |
| Arousal index, events/h | 243 | 10.99 | 5.63 |
| WASO, % | 240 | 9.4% | 10.3% |
| Total sleep time, min | 243 | 486.69 | 78.32 |
| Stage N1 sleep, min | 204 | 44.20 | 23.51 |
| Stage N2 sleep, min | 204 | 259.16 | 55.14 |
| Stage N3 sleep, min | 204 | 72.16 | 36.87 |
| REM sleep, min | 204 | 106.39 | 34.80 |
| Early morning N3 sleep, % | 202 | 49.0% | 3.5%* |
| JESS in unmedicated conditions | 183 | 15.79 | 4.79 |
| MSFsc | 204 | 4:28 AM | 1 h 48min |

Data are presented as the mean and standard deviation (SD), except where indicated by an asterisk (\*), which denotes standard error of the mean (SE). MSLT: multiple sleep latency test; BMI: body mass index; SOREMP: sleep onset rapid eye movement period; PSG: polysomnography; PLMI: periodic limb movement; WASO: wake after sleep onset; REM: rapid eye movement; Early morning N3 sleep: N3 within 2 h prior to the scheduled wake time; JESS: Japanese version of the Epworth Sleepiness Scale; MSFsc: sleep-corrected mid-sleep on free days.

### PRS Analysis

The primary objective of this study was to classify subtypes of IH, a heterogeneous disorder, using PRSs derived from various sleep-related phenotypes. The target set comprised IH patients with genome-wide genotype data as previously described. Base sets for sleep traits related to “sleep measurement (EFO_0004870)” and “sleep disorders (EFO_0008568)” were obtained from the PGS Catalog(*13, 14*). These included PRSs for “chronotype (morningness/eveningness)”, “sleep duration”, “sleeplessness/insomnia”, and “daytime napping and dozing”, all of which are registered in the catalog and were primarily derived from genetic studies conducted using the UK Biobank. The definitions of sleep traits are shown in Supplementary Table S3. The four abovementioned traits were used for the PRS analysis.

Given that methods for calculating PRSs can yield varying results depending on the approach used, we incorporated multiple calculation methods to enhance the robustness of our findings. From Tanigawa et al.(*19*), a single PRS method was available and thus adopted without modification. We utilized data from Privé et al.(*20*), Weissbrod et al.(*21*), and Ma et al.(*22*), each of which evaluated several PRS calculation methods and highlighted the strengths and limitations of each approach. In line with their methodologies, we selected the PRS with the highest incremental R² value or other relevant performance metrics (e.g., <u>AUC</u>) as the base data for our analysis. This approach ensured the selection of the most predictive PRS for the target phenotype while also accounting for variability across methods. Building on this approach, from the PGS Catalog, the PGS001055, PGS001001, PGS002209, and PGS002684 base sets were used for “chronotype (morningness/eveningness)”; PGS001150 and PGS002196 were used for “sleep duration”; PGS001932 and PGS003328 were used for “sleeplessness/insomnia”; and PGS000926, PGS002212, and PGS001000 were used for “daytime napping and dozing”.

The pgsc_calc workflow was utilized to calculate PRSs for each subject using scoring files published in the PGS Catalog(*13*). This workflow relies on the open-source software PLINK 2(*23*), PGS Catalog Utilities(*13*), and FRAPOSA(*13*). Genetic ancestry was assessed through a PCA implemented in the pgsc_calc pipeline (v2.0.0), with the 1000 Genomes Project serving as the reference panel. The calculated principal components (PCs) were used to adjust and contextualize PRSs for genetic ancestry.

PRSs were calculated in the IH target set for each sleep trait using multiple base sets (PGS identifiers), as detailed above. Multiple PRSs were thus generated for each trait. To identify individuals in the extreme tails of the PRS distributions (top or bottom 0.5%, 1%, 2.5% and 5%), the following approach was adopted: if an individual fell into the extreme group (upper or lower percentile) in any of the PGS-derived PRS distributions for a given sleep trait, they were classified as belonging to the extreme group for that trait, regardless of their percentile status in the other PGS-derived PRS distributions (Supplementary Figure S1). Subsequently, the numbers of IH patients and controls were determined for each extreme group. Frequencies between the two groups were compared using odds ratios (ORs) and either Fisher’s exact test or the chi-square test. Logistic regression analysis was used to compare IH patients and controls within the extreme PRS groups, with group status (IH patient or control) as the dependent variable and PRS group (top vs bottom percentile) as the independent variable. In addition, a logistic regression analysis was performed to assess group-wise differences in PRS values across the full distribution. As previously described, several base datasets were available for each sleep trait. To integrate multiple PRSs calculated for a single sleep trait into a representative value per sample, PC1 was calculated from the PRSs derived from these base datasets. The logistic regression analysis was then conducted with group status (IH patient or control) as the dependent variable, PC1 as the independent variable, and sex and array as covariates. Statistical significance was adjusted using the Bonferroni correction to account for multiple comparisons, based on three extreme percentiles (0.5%, 1%, 2.5% and 5%) and four sleep traits (total of 12 comparisons), resulting in an adjusted threshold of *P*<3.1×10^−3^ (0.05/16). Statistical analyses were performed using R software (v 4.3.0).

Clinical data were compared between the top and bottom 2.5% (or 5%) of IH patients stratified by PRS analysis as described below. Data were compared by linear regression analysis, with clinical data as the dependent variable, group status (top or bottom) as the independent variable, and sex, age, 24-h PSG availability, and body mass index (BMI) as covariates. The Bonferroni correction was used to control for multiple testing across 16 clinical measures, setting the corrected significance level at *P*<3.1×10^−3^ (0.05/16).

### MR analysis

A two-sample MR analysis was conducted using the inverse-variance weighted (IVW) approach(*24*) as the primary analysis, with MR-Egger regression(*25*), the weighted median method(*26*), and the corrected MR-PRESSO approach(*27*) as sensitivity analyses. Outcome data were derived from a GWAS of IH, based on the imputed genotype data (IH n = 303, controls n = 2918), as detailed in the Subjects, Genotyping, and Imputation section. Two sets of exposure data associated with chronotype were obtained from GWAS summary statistics of the UK Biobank (IEU OpenGWAS project: ukb-b-4956) and 23andMe(*28*). Chronotype was defined as indicated in Supplementary Table S3 for the UK Biobank and Supplementary Table S4 for 23andMe. The exposure datasets were mainly derived from individuals of European ancestry. When selecting genetic variants as instruments for the exposure, a significance threshold of *P*<6.0×10^−9^ (based on permutation testing reported in a chronotype GWAS(*28*)) was applied as a stringent threshold. Genetic variants that were in low linkage disequilibrium (*r²*<0.001) within ±250 kb, had a MAF >5% in our IH GWAS, and were non-palindromic were included. A total of 76 and 37 genetic variants associated with chronotype from the UK Biobank (ukb-b-4956) and 23andMe datasets, respectively, were selected as instruments. In addition, a genome-wide significance threshold of *P*<5.0×10^−8^ was also employed as the conventional threshold(*29*). The same selection criteria were applied, resulting in 106 and 50 genetic variants retained as instruments from the UK Biobank (ukb-b-4956) and 23andMe datasets, respectively. The full list of selected genetic variants used as instruments is provided in Supplementary Tables S5 and S6 for the UK Biobank (ukb-b-4956) and 23andMe datasets, respectively. Effect sizes (beta) and standard errors (SEs) for the instrumental variables were standardized to express the associations per one standard deviation (SD) increase in exposure, enabling effect estimates to be interpreted in SD units. To assess instrument strength, the F-statistic for each genetic variant was calculated as the squared association estimate between the variant and the exposure, divided by the squared SE. The mean F-statistic across all instruments was then computed. We also evaluated potential violations of the exclusion restriction assumption, including horizontal pleiotropy, using Cochran’s Q test, the MR-Egger intercept test, and the MR-PRESSO global test. All MR analyses were conducted using the TwoSampleMR (v 0.6.9) and MR-PRESSO (v 1.0) packages in R (v 4.3.0). Statistical significance was set at *P*<0.05 for all tests.

Given the clinical heterogeneity of IH, a subgroup MR analysis stratified by diagnostic method was conducted to determine whether the causal effect differed between IH patients with objectively confirmed long sleep duration (n = 57) (≥660 min on 24-h PSG) and those who were diagnosed using MSLT (n = 246). To compare the causal estimates between the 660-min PSG and MSLT groups, a two-sided Z-test was performed using the β-values and corresponding SEs derived from the MR analyses.

## Results

### PRS Analysis

We constructed a target set of genome-wide SNP data for 303 IH patients and 2918 controls using SNP arrays and genotype imputation. Base sets of sleep traits related to “sleep measurement (EFO_0004870)” and “sleep disorders (EFO_0008568)” were obtained from the PGS Catalog(*13, 14*). The base sets included scores for “chronotype (morningness/eveningness)”, “sleep duration”, “sleeplessness/insomnia”, and “daytime napping and dozing”, as registered in the PGS Catalog. Using these base data, PRSs of each sleep trait in the target data were then calculated with PCA-based adjustment. Statistical significance was assessed using Bonferroni correction for multiple comparisons, resulting in an adjusted threshold of *P*<3.1×10^−3^. Individuals in the top and bottom percentiles (0.5%, 1%, 2.5% and 5%) of each PRS distribution were identified. Further methodological details are provided in the Subjects and Methods section.

IH patients were significantly overrepresented in the top PRS percentiles for chronotype (eveningness) and daytime napping/dozing (Figure 1 and Supplementary Table S7). With regard to chronotype, IH patients were more prevalent in all top PRS percentiles. The associations remained statistically significant after Bonferroni correction in the top 0.5% (*P* = 3.9×10^−5^, OR = 3.40), top 1% (*P* = 1.9×10^−4^, OR = 2.48) and top 5% (*P* = 5.5×10^−5^, OR = 1.81) groups. The association in the top 2.5% group was nominally significant (*P* = 3.9×10^−3^, OR = 1.71) but did not survive multiple testing correction. Conversely, in the bottom 2.5% percentile (morningness), IH patients were significantly underrepresented (bottom 2.5%: *P* = 2.9×10^−3^, OR = 0.44; bottom 5%: *P* = 2.6×10^−3^, OR = 0.57). Similar findings were observed for daytime napping/dozing PRSs, in which IH patients were overrepresented in the top 0.5% (*P* = 1.6×10^−5^, OR = 3.80) group, with a trend at 1% (top 1%: *P* = 0.027, OR = 1.87). For both traits, the ORs were consistently >1 in the top percentiles and <1 in the bottom percentiles, reinforcing the association between PRS and disease status.

**Figure 1.**
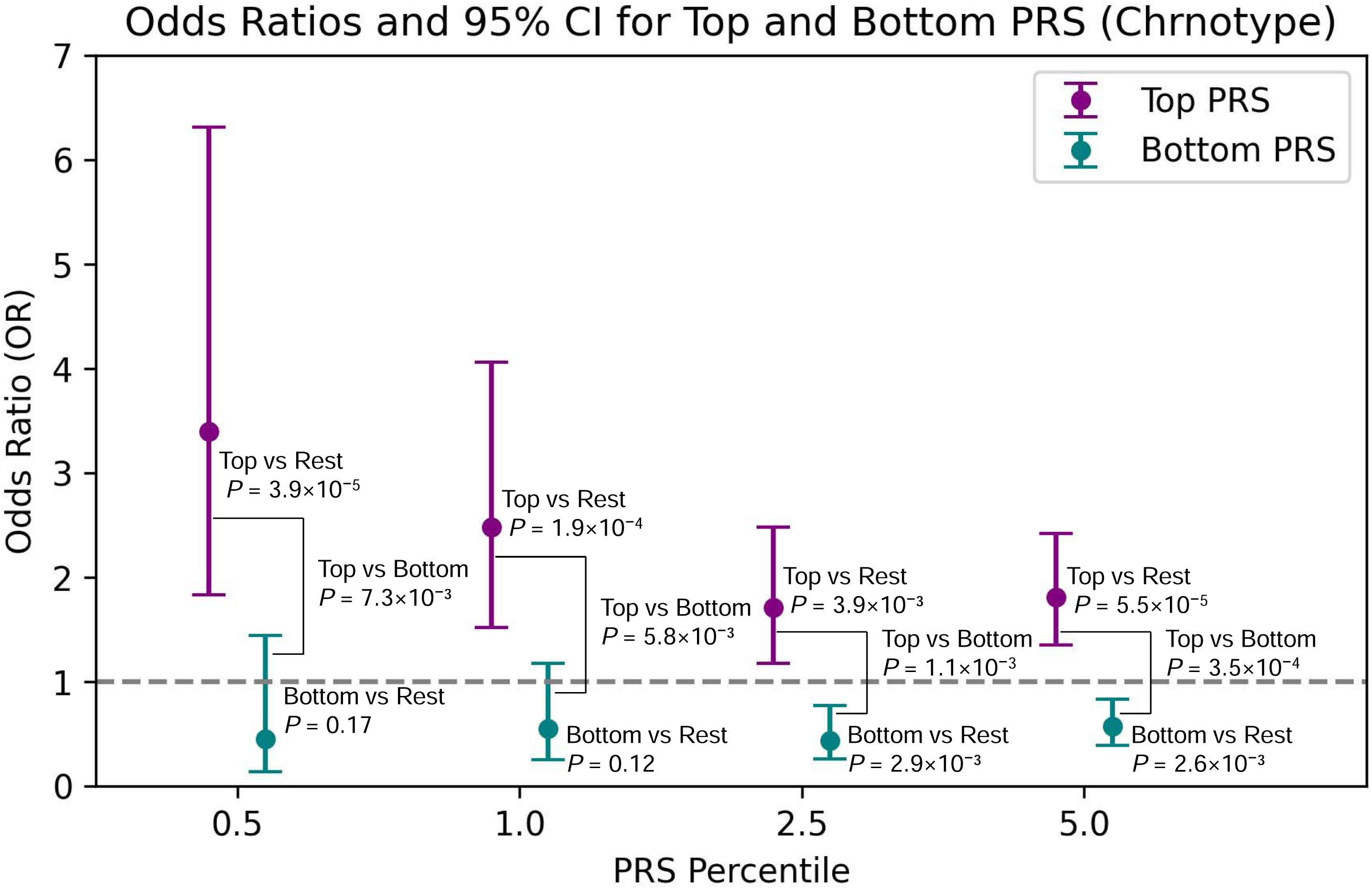
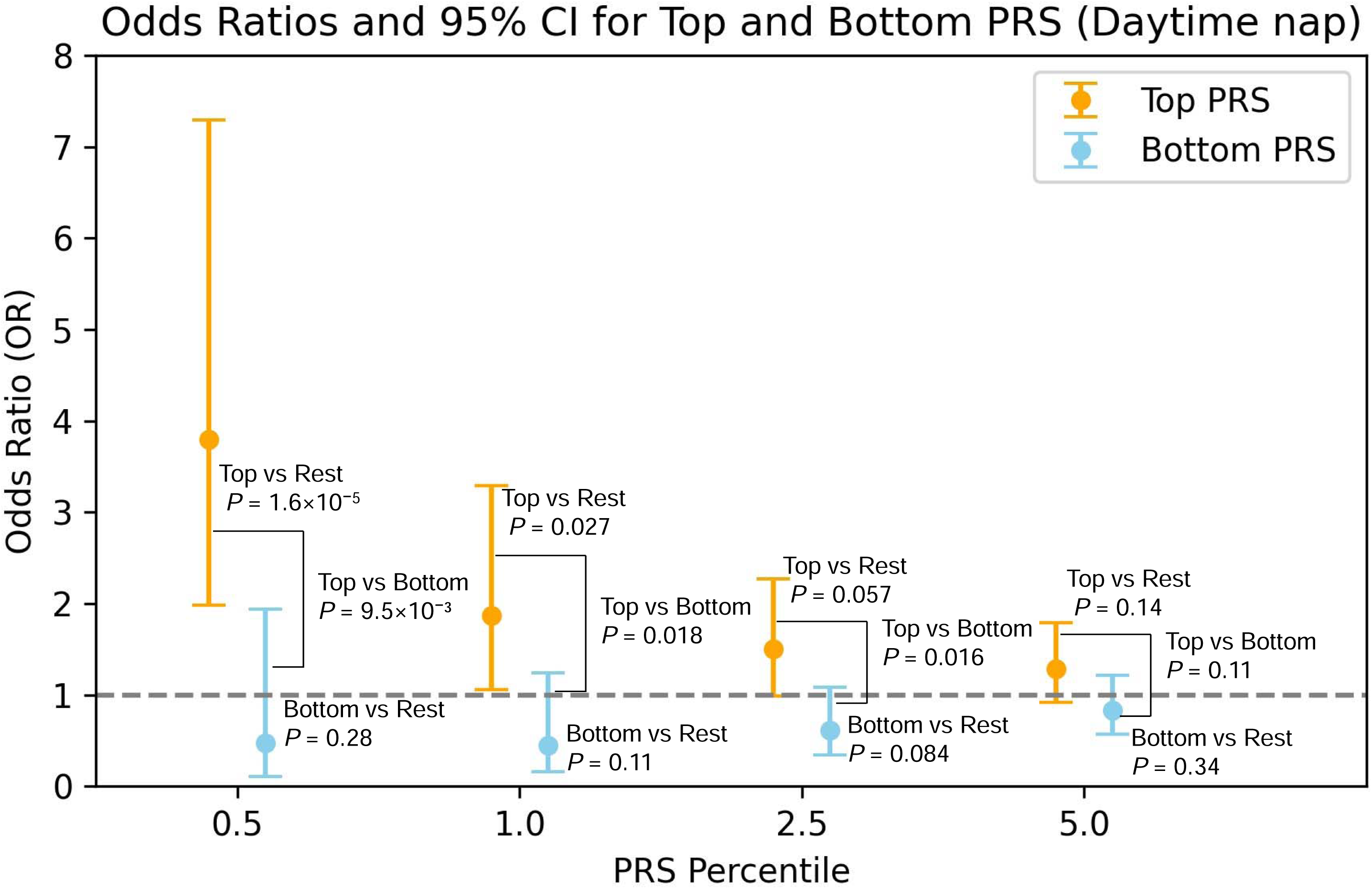
Polygenic risk score (PRS) associations with idiopathic hypersomnia (IH). Panel (A) shows odds ratios (ORs) and 95% confidence intervals (CIs) for IH in the top and bottom PRS percentiles for chronotype. Panel (B) shows ORs and 95% CIs for IH in the top and bottom PRS percentiles for daytime napping.

To further validate these findings, the top and bottom percentiles were directly compared. Despite relatively small sample sizes in these extreme groups, IH patients remained overrepresented among the top PRSs for chronotype (*P* = 7.3×10^−3^ at 0.5%, *P* = 5.8×10^−3^ at 1%, *P* = 1.1×10^−3^ at 2.5%, *P* = 3.5×10^−4^ at 5%) and daytime napping/dozing (*P* = 9.5×10^−3^ at 0.5%, *P* = 0.018 at 1%, *P* = 0.016 at 2.5%) (Figure 1 and Supplementary Table S7). Finally, group-wise comparisons of PRS-derived PC1 scores across the full distribution, conducted using logistic regression analysis, revealed significantly higher scores in IH patients for chronotype (*P* = 3.0×10^−5^, OR = 1.18), which remained statistically significant after Bonferroni correction (Supplementary Table S8). To visually illustrate the distributional difference in PC1 between the groups for chronotype, a boxplot was generated, and the difference was also assessed using the Wilcoxon rank-sum test (*P* = 2.9×10^−4^) (Figure 2). The association with daytime napping/dozing was nominally significant (*P* = 9.8×10^−3^, OR = 1.13) (Supplementary Table S8). No significant differences were found for PRSs for sleep duration or sleeplessness/insomnia in either the extreme or full-distribution comparisons (Supplementary Tables S7 and S8).

**Figure 2.**
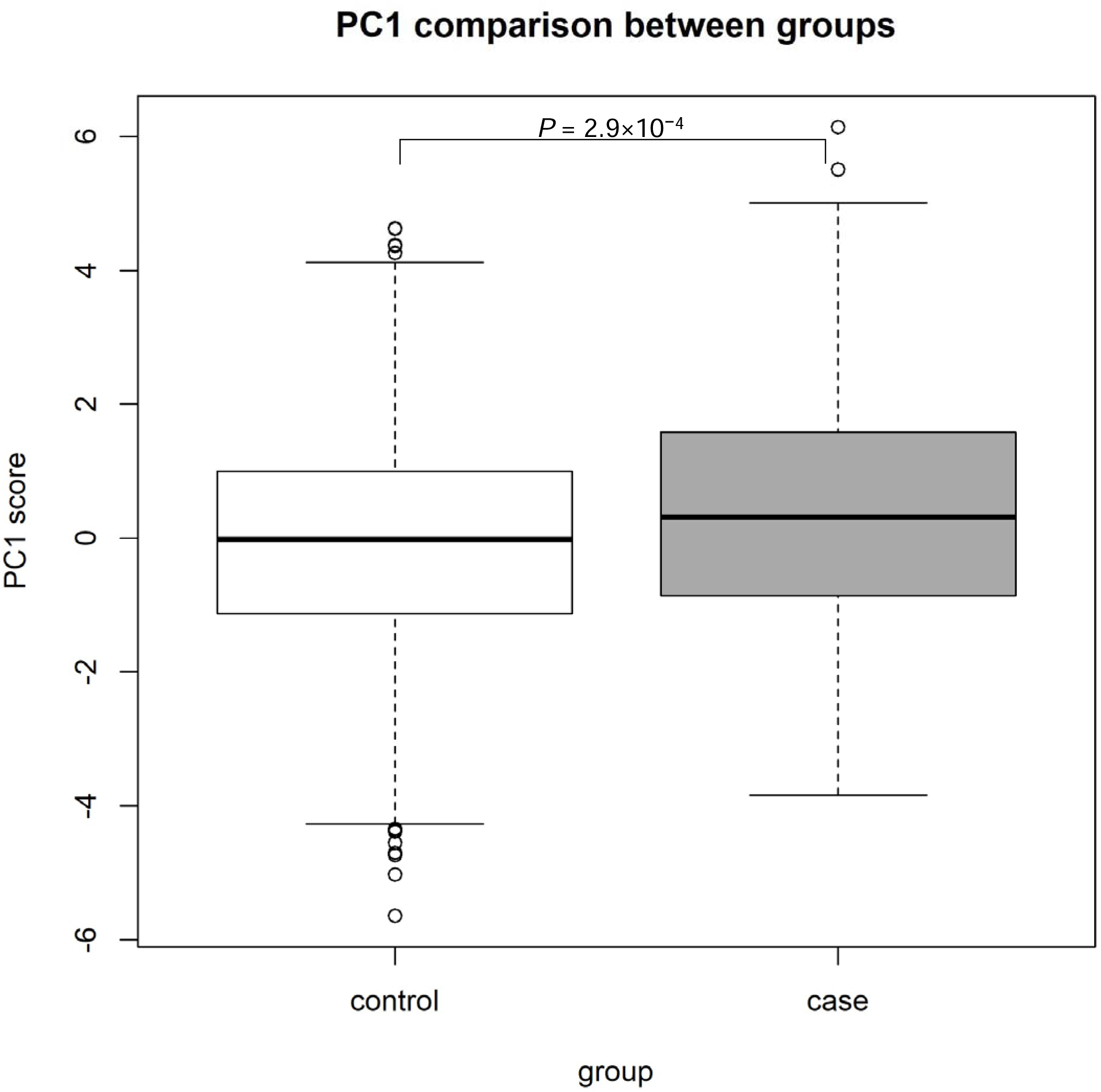
First principal component (PC1) distribution of chronotype polygenic risk scores (PRSs) in idiopathic hypersomnia (IH) patients and controls. Distribution of the PC1 derived from PRSs for chronotype in patients with IH and controls. PC1 was calculated using four PRSs (PGS001001, PGS001055, PGS002209, and PGS002684) and reflects the composite genetic tendency toward chronotype. Positive PC1 values indicate a genetic predisposition toward eveningness. The group difference was assessed using the Wilcoxon rank-sum test (*P* = 2.9×10□^4^).

### Phenotypic profiling of patients stratified using PRS analysis

We investigated whether IH patients with extreme PRS values for significant traits—specifically chronotype (morningness/eveningness) and daytime napping and dozing—exhibited distinct clinical characteristics. This analysis used objective sleep parameters derived using PSG and the MSLT, as well as subjective parameters including the MSFsc, and the JESS evaluated under unmedicated conditions. We also assessed the occurrence of N3 (slow-wave sleep) within 2 hours prior to the scheduled wake time, as a previous study showed that IH patients have more frequent N3 sleep episodes at the end of the night(*30*). When comparing the top and bottom 2.5% patient groups in terms of chronotype, N3 sleep occurring 2 hours before wake time, a pattern considered a characteristic feature of IH, was more frequently observed in the bottom group (morningness) than in the top group (eveningness). Although this difference did not reach statistical significance after Bonferroni correction (*P* = 7.4×10^−3^), the frequency was notably higher in the bottom group (91.7%) compared to the top group (48.0%) (Table 2). When compared with the remaining IH patients in the study (48.0%), the proportion of individuals showing this early morning N3 sleep pattern was not significantly different from the top group (*P* = 0.78), but was lower compared with the bottom group (*P* = 0.016). A similar trend was observed in the comparison of the top and bottom 5% patient groups based on chronotype, with the early morning N3 sleep pattern occurring more frequently in the bottom group than in the top group (*P* = 0.030) (Supplementary Table S9). Interestingly, there was no significant difference in MSFsc between the top and bottom groups, even though MSFsc is an established subjective measure of chronotype. No other significant differences were found in the remaining clinical or sleep parameters, either in the extreme chronotype groups or in the extreme daytime napping/dozing PRS groups (Table 2 and Supplementary Tables S9, S10 and S11).

**Table 2.** Comparison of sleep-related traits in IH patients with extreme PRS values for eveningness and morningness chronotypes (top and bottom 2.5%).

| Variable | Eve PRS:<br>Mean | Eve PRS:<br>SD (or SE*) | Mor PRS:<br>Mean | Mor PRS:<br>SD (or SE*) | *P* (Eve vs.<br>Mor) | Eve PRS:<br>N | Mor PRS:<br>N | *P* (Eve vs.<br>Others) | *P* (Mor vs.<br>Others) |
| --- | --- | --- | --- | --- | --- | --- | --- | --- | --- |
| Mean sleep latency (MSLT), min | 5.94 | 4.24 | 5.15 | 5.43 | 0.64 | 29 | 14 | 0.50 | 0.26 |
| SOREMP (MSLT), number | 0.34 | 0.48 | 0.36 | 0.50 | 0.84 | 29 | 14 | 0.56 | 0.69 |
| Sleep efficiency (PSG), % | 86.2% | 9.7% | 91.5% | 7.8% | 0.12 | 30 | 12 | 0.30 | 0.36 |
| Apnea hypopnea index (PSG),<br>events/h | 2.66 | 3.05 | 1.08 | 0.74 | 0.07 | 30 | 12 | 0.39 | 0.07 |
| Sleep latency (PSG), min | 14.13 | 15.94 | 12.92 | 22.03 | 0.91 | 30 | 12 | 0.64 | 0.56 |
| PLMI (PSG), events/h | 1.40 | 2.78 | 0.75 | 1.56 | 0.46 | 29 | 12 | 0.20 | 0.80 |
| Arousal index (PSG), events/h | 10.72 | 5.02 | 9.51 | 2.79 | 0.35 | 29 | 12 | 0.89 | 0.39 |
| WASO (PSG), % | 11.6% | 9.4% | 7.4% | 8.1% | 0.23 | 29 | 12 | 0.32 | 0.58 |
| Total sleep time (PSG), min | 491.84 | 91.28 | 505.00 | 55.20 | 0.74 | 29 | 12 | 0.60 | 0.46 |
| Stage N1 sleep (PSG), min | 47.69 | 30.67 | 38.38 | 11.20 | 0.31 | 26 | 12 | 0.37 | 0.39 |
| Stage N2 sleep (PSG), min | 264.75 | 64.25 | 271.13 | 44.30 | 0.82 | 26 | 12 | 0.54 | 0.48 |
| Stage N3 sleep (PSG), min | 78.02 | 34.67 | 78.79 | 40.26 | 0.82 | 26 | 12 | 0.41 | 0.57 |
| REM sleep (PSG), min | 103.42 | 46.05 | 116.71 | 29.21 | 0.33 | 26 | 12 | 0.70 | 0.35 |
| Early morning N3 sleep (PSG), % | 48.0% | 10.2%* | 91.7% | 8.3%* | 0.0074 | 25 | 12 | 0.78 | 0.016 |
| JESS in unmedicated conditions | 14.55 | 5.50 | 15.45 | 5.65 | 0.58 | 20 | 11 | 0.23 | 0.91 |
| MSFsc | 4 h 41 min | 1 h 50 min | 4 h 36 min | 2 h 7 min | 0.98 | 26 | 12 | 0.61 | 0.91 |

Data are presented as the mean and standard deviation (SD), except where indicated by an asterisk (\*), which denotes standard error of the mean (SE).
Eve: eveningness; Mor: morningness; MSLT: multiple sleep latency test; SOREMP: sleep onset rapid eye movement period; PSG: polysomnography; PLMI: periodic limb movement; WASO: wake after sleep onset; REM: rapid eye movement; Early morning N3 sleep: N3 within 2 hours prior to the scheduled wake time; JESS: Japanese version of the Epworth Sleepiness Scale; MSFsc: sleep-corrected mid-sleep on free days.

### MR analysis

Robust associations between eveningness-related PRSs and IH were observed in both the full-distribution and extreme-percentile analyses. These findings prompted us to explore a potential causal relationship using two-sample MR analysis. Significant genetic variants associated with chronotype (*P*<6.0×10^−9^) from a GWAS of the UK Biobank (ukb-b-4956) were used as exposure data. The outcome data were obtained from a GWAS of IH, which was also employed in the PRS analysis described above. IVW analysis with random effects revealed a causal relationship between eveningness and IH (*P* = 1.3×10^−3^, β = 0.042) (Figure 3, and Supplementary Table S12). Sensitivity analyses using MR-Egger regression, the weighted median method, and the corrected MR-PRESSO approach supported the observed causal effect. We also performed MR analysis using associated genetic variants from a chronotype GWAS of 23andMe as a replication study. The significant causal effect of eveningness on IH was replicated in this independent dataset using the IVW approach (*P* = 0.037, β = 0.036), with consistent results in sensitivity analyses.

**Figure 3.**
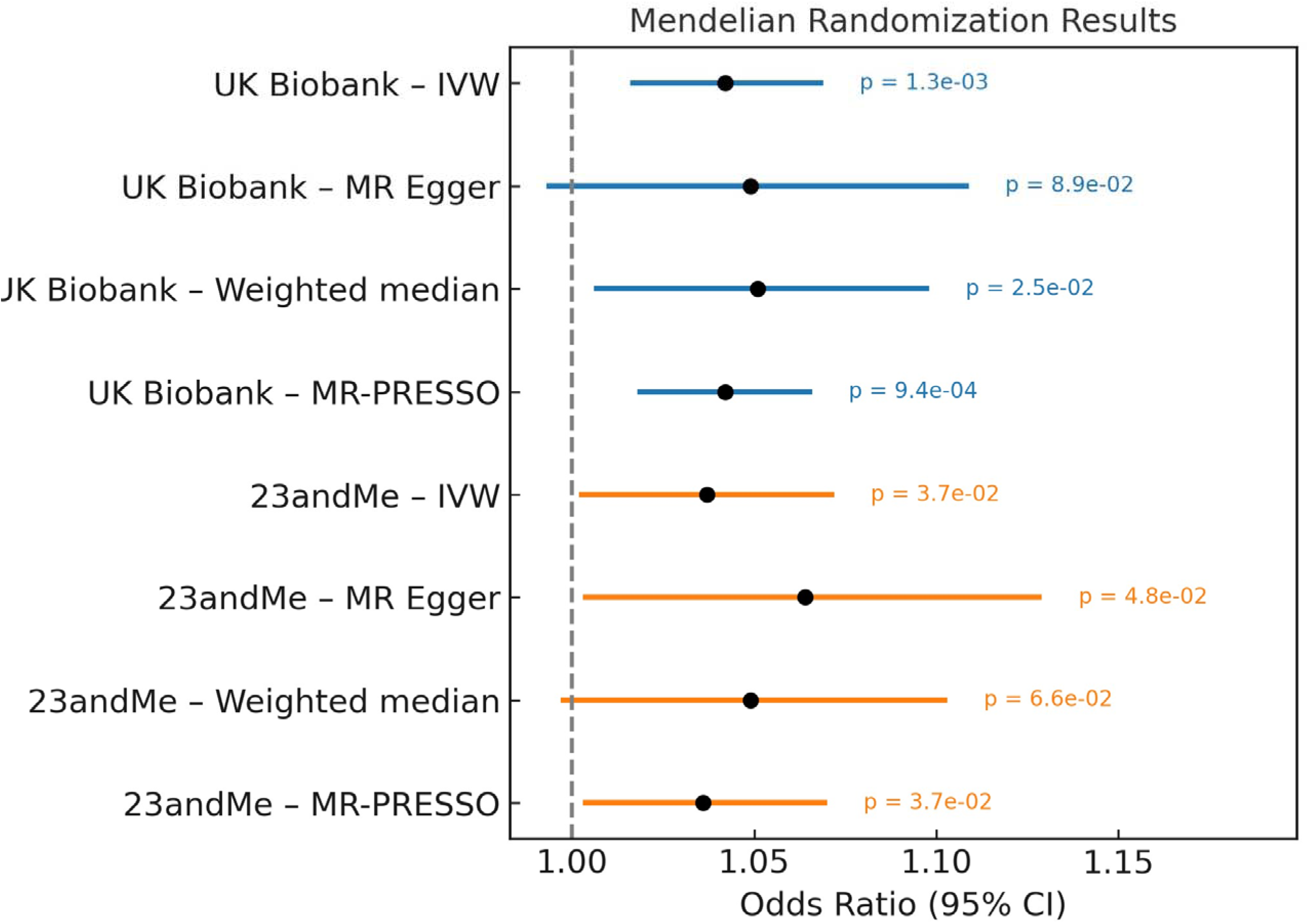
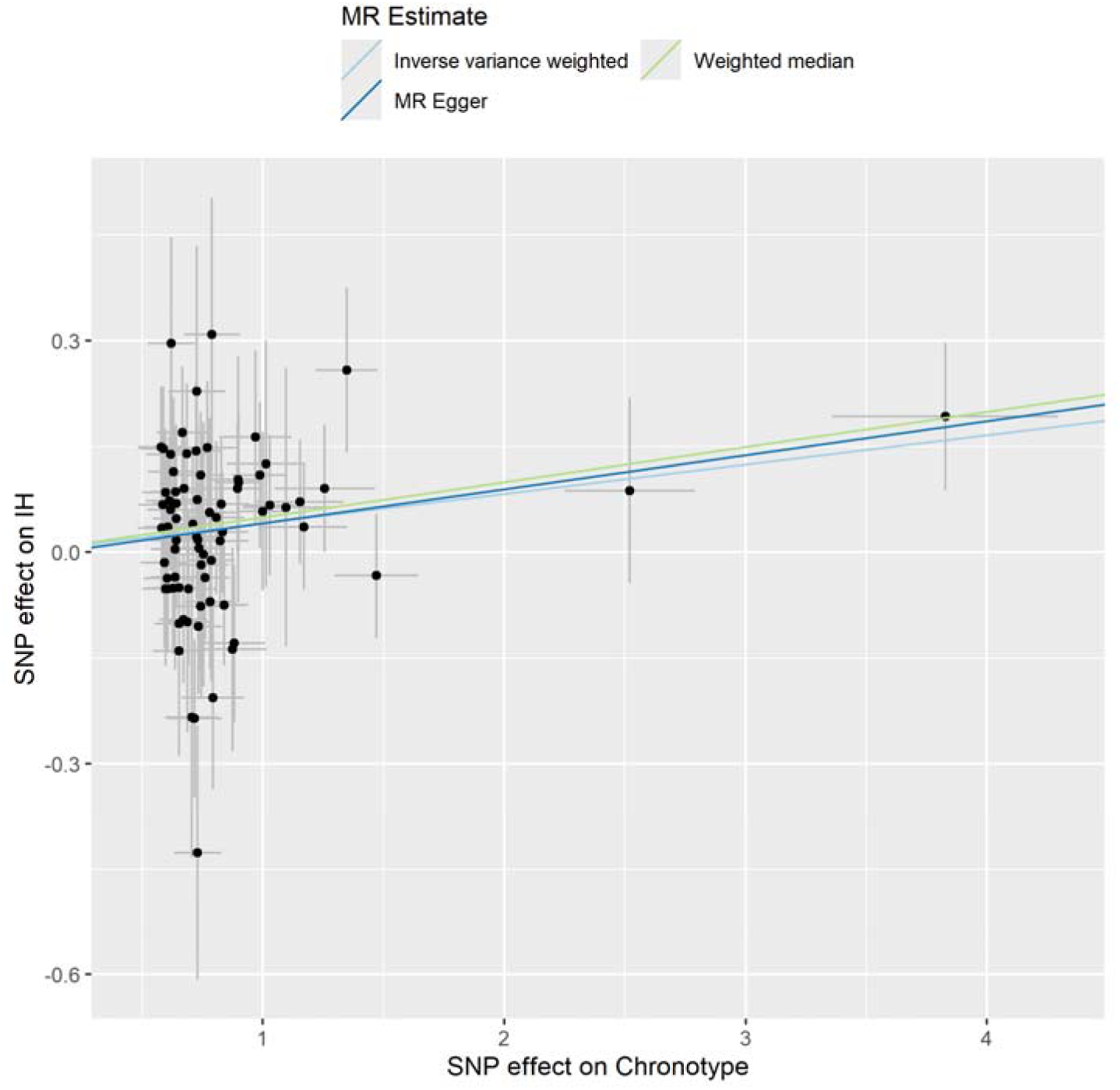
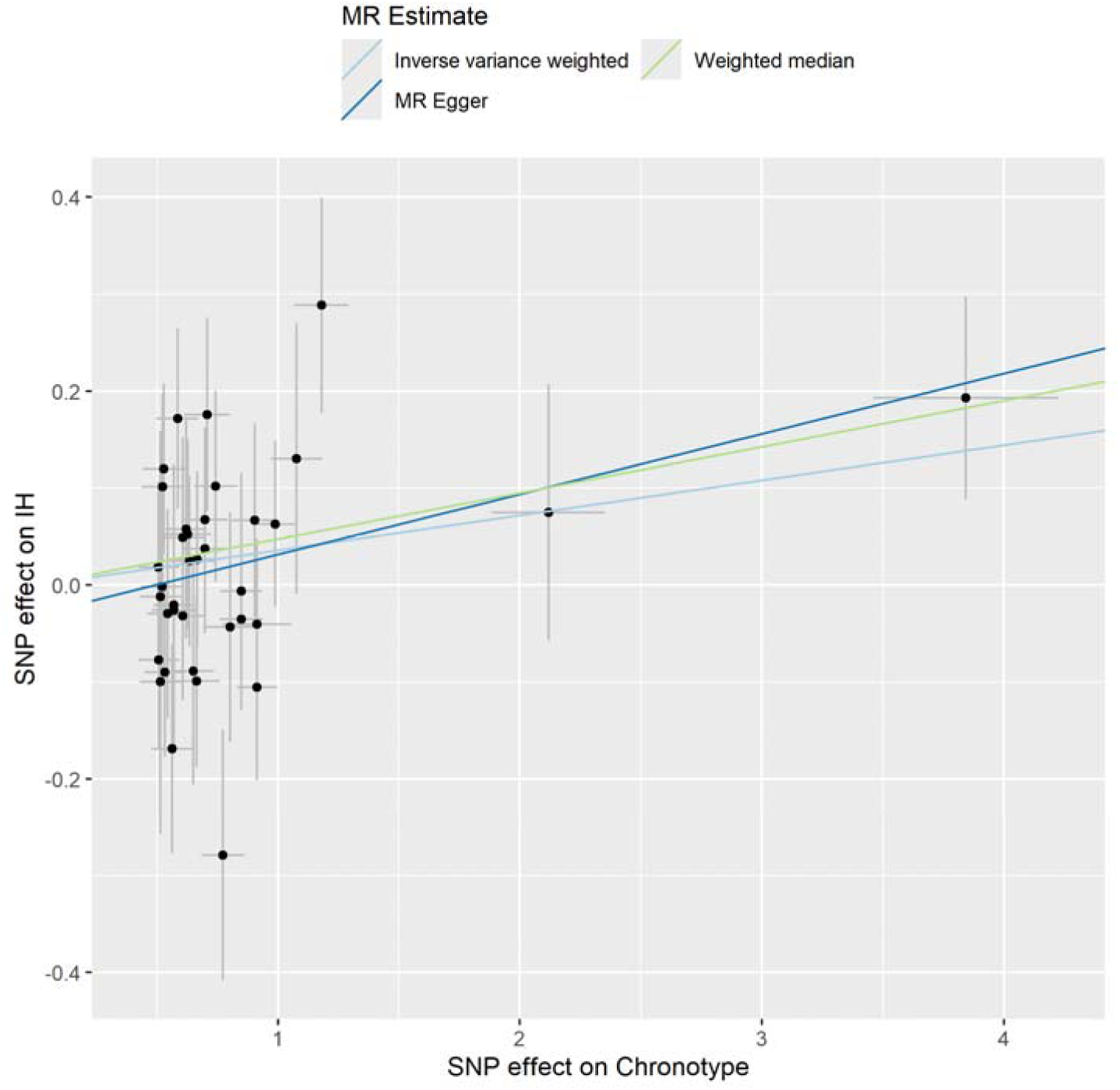
Forest plot and scatter plots showing Mendelian randomization (MR) estimates for the effect of chronotype on idiopathic hypersomnia (IH) using various MR methods with genetic instruments selected at *P* < 6×10^−9^. Panel (A) shows a forest plot generated using various MR methods. The estimated effects, expressed as odds ratios (ORs), are shown per one standard deviation (SD) increase in the exposure along with 95% confidence intervals (CIs). A positive β-value indicates a genetic tendency toward eveningness. Thus, an OR >1 suggests that eveningness is associated with an increased risk of IH. Each row in the forest plot corresponds to a different MR method, each of which utilizes a distinct set of genetic instruments associated with chronotype. Panels (B) and (C) display MR scatter plots of IH risk against chronotype exposure. Each dot represents a genetic variant, with the effect on eveningness (x-axis) plotted against the effect on IH risk (y-axis, in log odds), standardized in SD units. Regression lines show causal estimates from different MR methods; a positive slope indicates that a genetic tendency toward eveningness is causally associated with higher IH risk. Panel (B) used data from the UK Biobank, and Panel (C) used data from 23andMe. Effect sizes and standard errors (SEs) for chronotype were standardized; error bars represent SE values.

To assess the robustness and validity of the MR findings, we examined the strength of the instruments and evaluated key assumptions, including heterogeneity and directional pleiotropy. The mean *F*-statistic of the genetic instruments for chronotype from the UK Biobank and 23andMe was 49.88 and 59.03, respectively, suggesting that the instruments were sufficiently strong (*F*>10) (Supplementary Table S12). Cochran’s *Q* test showed no significant heterogeneity in the IVW analysis estimates (*P* = 0.71 for UK Biobank; *P* = 0.60 for 23andMe). Furthermore, there was no evidence of directional pleiotropy, as indicated by non-significant MR-Egger intercepts (*P* = 0.79 for UK Biobank; *P* = 0.30 for 23andMe).

To further assess the robustness of the causal inference, we performed an additional MR analysis using the conventional threshold for instrument selection (*P*<5.0×10^−8^). The results remained consistent with those obtained using the stringent threshold (*P*<6.0×10^−9^), suggesting the robustness of the association (*P* = 0.010, β = 0.030 for UK Biobank; *P* = 0.031, β = 0.036 for 23andMe) (Supplementary Figures S2 and S3, and Supplementary Table S12). The robustness and validity of these MR findings were confirmed, and the findings remained robust even under a more stringent instrument selection threshold, reinforcing the credibility of a causal relationship between eveningness and IH.

To account for the clinical heterogeneity of IH, we conducted a subgroup MR analysis stratified by the availability of 24-h PSG data. Specifically, patients who underwent 24-hour PSG and exhibited ≥660 min of total sleep time (n = 57) were compared with those who were diagnosed using standard overnight PSG and MSLT (n = 246). The ≥660-min 24-hour PSG group represents objectively confirmed long sleep duration and is therefore more likely to reflect true IH cases, whereas the standard overnight PSG and MSLT group may include individuals with unrecognized sleep deprivation or phenocopies. This subgroup analysis assessed whether the causal estimates from MR analyses differed between the two diagnostic subgroups. The results showed consistent effect directions across both subgroups. Although the effect size appeared larger in the ≥660-min 24-h PSG group, a two-sided Z-test comparing the IVW analysis estimates revealed no significant difference (Table 3). Sensitivity analyses using MR-Egger, weighted median, and MR-PRESSO methods yielded similar effect directions and magnitudes (Supplementary Table S13), suggested a consistent trend across diagnostic subgroups.

**Table 3.** Comparisons of MR results (inverse-variance weighted [IVW] method) for chronotype exposure and IH defined by 24-h PSG (≥660 min) and MSLT.

| Exposure | 24hPSG_IH (n = 57) |  |  | MSLT_IH (n = 246) |  |  | Z<sup>2</sup> | P (z-test) |
| --- | --- | --- | --- | --- | --- | --- | --- | --- |
|  | Beta | SE | P | Beta | SE | P |  |  |
| UKB ukb-b-4956<br>(P<5×10<sup>-8</sup>) | 0.045 | 0.027 | 0.093 | 0.025 | 0.013 | 0.050 | 0.66 | 0.51 |
| 23andMe<br>(P<5×10<sup>-8</sup>) | 0.078 | 0.038 | 0.040 | 0.022 | 0.018 | 0.22 | 1.33 | 0.18 |
| UKB ukb-b-4956<br>(P<6×10<sup>-9</sup>) | 0.053 | 0.032 | 0.093 | 0.040 | 0.015 | 0.0083 | 0.37 | 0.71 |
| 23andMe<br>(P<6×10<sup>-9</sup>) | 0.079 | 0.043 | 0.063 | 0.025 | 0.020 | 0.22 | 1.15 | 0.25 |

Beta and standard errors (SE) values are presented.
PSG: polysomnography; MSLT: multiple sleep latency test; UKB: UK Biobank.

## Discussion

We performed a PRS analysis as the base set of sleep traits and target set of IH to stratify IH patients from the perspective of genetic sleep traits. IH patients with a greater genetic predisposition to eveningness or to daytime napping and dozing were overrepresented in the top PRS percentiles, indicating that stratification based on genetic sleep profiles is feasible. Remarkably, the ORs for IH among individuals in the top PRS percentiles exceeded 2.0, reaching 3.40 in the top 0.5% and 2.48 in the top 1.0% for chronotype-related PRSs and 3.80 in the top 0.5% for daytime napping and dozing (Figure 1 and Supplementary Table S7), suggesting PRS-based stratification is robust and has potential clinical utility.

IH is considered a heterogeneous sleep disorder. A subset of IH patients exhibited extreme PRSs for eveningness or for daytime napping and dozing, suggesting that IH may involve distinct underlying etiologies that vary between individuals. Interestingly, no significant differences were observed in MSFsc, a widely used subjective measure of chronotype, among patients with extreme eveningness PRSs (4.41 h), morningness PRSs (4.36 h), or the overall IH group (4.28 h) (Tables 1 and 2). The reported average MSFsc in the general Japanese population is approximately 4.31 h, which does not substantially differ with that of IH patients(*31*). These results suggest that sleep regulation per se may be disrupted after the onset of IH, potentially limiting the reliability of MSFsc as an indicator of an individual’s underlying chronotype. It is also possible that eveningness acts as a predisposing factor in the pathogenesis of IH, but once the disease manifests, differences in chronotype between patients may become less apparent. A previous *in vitro* study using fibroblasts from IH patients and healthy controls reported attenuated amplitudes of core clock gene expression in IH patients(*32*). Therefore, future studies should investigate whether IH patients with extreme chronotype-related PRSs also exhibit altered clock gene dynamics in peripheral tissues and cells, such as fibroblasts.

A core symptom of IH is prolonged sleep, with patients often requiring more than 11 h of sleep per day(*30, 33*). Interestingly, in our study, neither individuals with extreme PRSs for sleep duration nor the overall PRS distribution for this trait showed a significant association with IH. This finding suggests that it may not be prolonged sleep per se, as a normative sleep phenotype, but rather other factors, such as eveningness, identified in this study, that contribute more critically to the development of IH.

We also explored whether individuals with IH who exhibit extreme PRSs for key sleep-related traits exhibit distinct clinical features. While genome-wide analyses revealed robust associations between PRS and IH susceptibility, the aim of this stratified phenotypic profiling based on PRS extremes was to determine whether genetic liability is reflected in observable clinical characteristics. N3 sleep occurring within 2 h prior to the PSG-defined wake time was more prevalent in the bottom PRS group (morningness) in the chronotype analysis (Table 2). Sleep drunkenness after awakening is a symptom of IH(*3, 30, 34, 35*). The potential association between the presence of deep sleep (N3) near wake time and sleep drunkenness remains an area of interest. Previous studies based on sleep examination findings reported that slow-wave sleep near the time of waking is no more common in individuals with sleep drunkenness than in those without(*3, 30*). Interestingly, although previous studies based on sleep phenotyping have suggested that individuals with an eveningness chronotype are more prone to sleep drunkenness(*3, 34*), our genetic analysis revealed that IH patients with a genetic predisposition toward morningness (i.e., bottom PRS group) were more likely to exhibit early morning N3 sleep. At first glance, this might appear to be a paradoxical association. However, it is also possible that the occurrence of N3 sleep near wake time and the manifestation of sleep drunkenness are not directly linked but rather represent distinct physiological phenomena. This distinction warrants further investigation to clarify the respective roles of N3 sleep near wake time and sleep drunkenness in the pathophysiology of IH. This relationship may suggest the presence of a pathophysiological mechanism in IH that is independent of eveningness-related genetic predisposition. The presence of N3 sleep shortly before the scheduled wake time may represent a biologically meaningful feature of IH. Although previous studies have reported that IH patients exhibit decreased N3 and increased N2 sleep overall(*36*), we observed no significant differences in N2 or total N3 sleep time between the two PRS-defined groups in our analysis. This observation raises the possibility that distinct subtypes of IH exist, with different underlying mechanisms, some related to circadian predisposition and others potentially reflecting dysregulation of homeostatic sleep pressure or arousal systems. Although further studies are needed to establish the clinical relevance of this pattern, it could potentially serve as a phenotypic marker for subclassifying IH patients, especially in relation to underlying genetic risk profiles.

MR analysis supported a causal relationship, identifying eveningness as a risk factor for the development of IH at the population level, beyond individuals with extreme PRSs (Figure 3). A previous cross-sectional study reported that IH patients are more frequently of the evening type and tend to be more alert in the evening than the morning(*34*). Our study thus provides genetic and causal evidence that substantiates these prior clinical observations and thus contributes to a better understanding of the pathophysiology of IH. By contrast, daytime napping and dozing was not subjected to MR analysis, as this trait represents a core symptom of IH rather than an independent risk factor. Although the effect sizes observed in MR analyses were more modest—likely due to its continuous, population-level nature—the consistent direction of association across both PRS and MR analyses reinforces the credibility of the link between eveningness and IH. These findings, further validated by sensitivity tests and replication in an independent cohort, provide strong evidence for a robust and potentially causal contribution of the eveningness chronotype to the pathogenesis of IH.

The subgroup MR analysis stratified by diagnostic method revealed consistent effect directions in both the ≥660-min 24-h PSG and standard PSG/MSLT groups, suggesting that the observed causal relationship between eveningness and IH is robust across diagnostic heterogeneity. Although the effect size appeared larger in the ≥660-min 24-h PSG group, the difference was not statistically significant (Table 3 and Supplementary Table S13). This finding supports the utility of 24-h PSG as a high-specificity diagnostic tool for identifying true IH cases(*1, 2*), while also reaffirming the clinical validity of MSLT-based diagnosis under ICSD-3-TR criteria(*4, 33*). Further studies incorporating more granular phenotyping, comprehensive genetic data, and larger sample sizes could clarify potential etiological distinctions between diagnostic subtypes of IH.

This study has several limitations. First, the number of individuals in the extreme PRS percentiles was relatively small, which may have reduced the statistical power for detecting phenotypic differences within stratified subgroups. Future studies with larger sample sizes or meta-analyses combining data from multiple cohorts may help improve statistical power and enable more robust subgroup analyses. Second, the generalizability of our findings may be limited. Our analysis was restricted to a Japanese population, and the PRSs used were constructed based on summary statistics predominantly derived from the UK Biobank, which primarily includes individuals of European ancestry. This ancestry mismatch is a well-recognized limitation in PRS analyses. In addition, cultural and demographic differences between the UK Biobank and our study population should be acknowledged. UK Biobank participants are predominantly older adults whereas our sample consists mainly of younger individuals (Table 1). Moreover, cultural norms and lifestyle factors that influence sleep behaviors may differ between British and Japanese individuals, potentially affecting the applicability of PRSs to different populations. Notably, Tanigawa et al.(*19*) reported that sleep trait PRSs constructed using UK Biobank data (white British) showed statistically significant predictive performance in East Asian populations (Supplementary Table S14). While we acknowledge that constructing sleep trait PRSs based on ancestrally matched populations would be ideal, the UK Biobank-based GWAS currently offers the largest available sample size for sleep-related traits. Therefore, we considered it the best available resource at this time and proceeded with our analyses accordingly. Third, although our MR approach supports a directional relationship between sleep traits and IH, we acknowledge that parental genotype effects could influence the observed associations. A within-family MR design would be necessary to fully disentangle these effects; however, such an analysis was beyond the scope of this study. Future studies employing within-family MR approaches may help clarify the role of parental genetic influences in the development of IH.

Collectively, the findings of this study not only shed light on the potential genetic underpinnings of IH but also underscore the importance of incorporating genetic sleep traits into future IH subtyping and treatment strategies. This integrative genomic approach may pave the way for personalized medicine in hypersomnia and contribute to a more refined understanding of its heterogeneous pathophysiology. Additionally, our findings suggest that an eveningness chronotype contributes to the risk of IH. This association raises the possibility that interventions targeting circadian phase or sleep-wake patterns could help modify disease risk or severity. However, further studies are needed to establish causal mechanisms and evaluate therapeutic efficacy.

## Data Availability

Genome-wide data supporting this study can be accessed upon application to NBDC Human Database (https://humandbs.biosciencedbc.jp/en/) (NBDC research ID: hum0264, Japanese Genotype-phenotype Archive (JGA) accession number: JGAS000508).

## Acknowledgements

The authors are deeply grateful to all participants in the present study. This study was supported by Grants-in-Aid for Scientific Research (B: 19H03588, 22H03006, and 23K24267) and (C: 21K07534 and 25K10826) from the Ministry of Education, Culture, Sports, Science and Technology of Japan, and by Grants-in-Aid for Multilayered Stress Diseases (JPMXP1323015483). The funders had no role in study design, data collection, analysis, the decision to publish, or preparation of the manuscript. The data used for this study (hum0082.v2 and hum0136.v3) were originally obtained by the Department of Human Genetics, Graduate School of Medicine, The University of Tokyo (led by Prof. Tokunaga), and the Genome Medical Science Project at the National Center for Global Health and Medicine (led by Dr. Mizokami). These datasets are available from the website of the NBDC Human Database (https://humandbs.dbcls.jp/en/) of the Database Center for Life Science (DBCLS) / the Joint Support-Center for Data Science Research (DS) of the Research Organization of Information and Systems (ROIS)(*37*).

## Author Contributions

T.M., M.S., T.K., K.T., and M.H. contributed to the conception and design of the study; T.M., M.S., K.T., N.K., T.K., T.K., Y.A., H.H., M.O., N.U., A.I., K.K., T.K., A.I., Y.K., A.H., Y.W., K.K., M.M., K.H., M.T., N.Y., M.O., N.O., H.K., T.K., Y.I., K.M., and M.H. contributed to acquisition and analysis of data; T.M., M.S., and M.H. contributed to drafting the text or preparing the figures.

