## supplemental tables and figs for "Polygenic risk scores and Mendelian randomization reveal circadian genetic contributions to idiopathic hypersomnia"

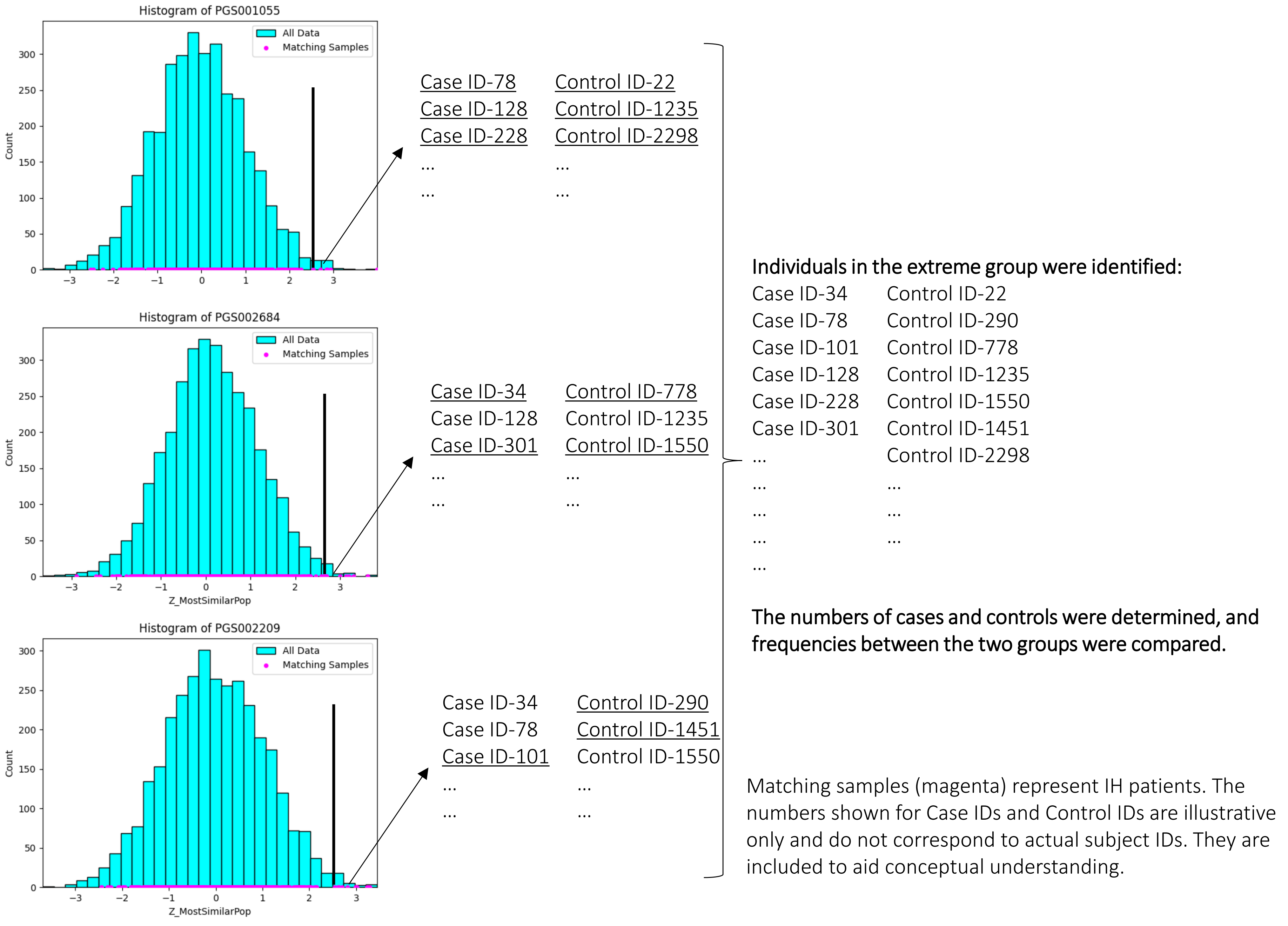


Supplementary Figure S1. Method for extracting individuals in the extreme PRS groups based on multiple PGS sources.

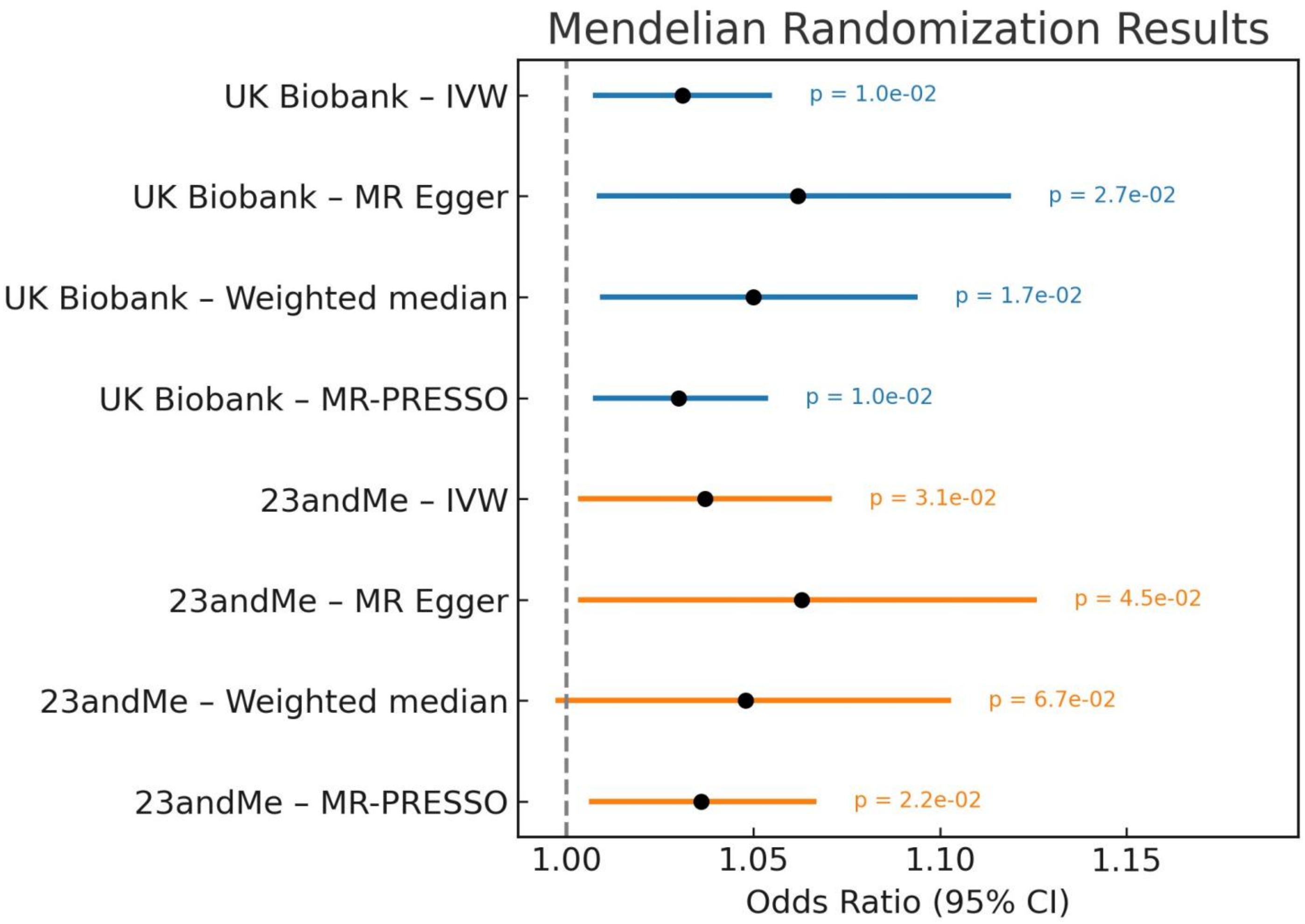


Supplementary Figure S2. Forest plot showing Mendelian randomization (MR) estimates for the effect of chronotype on idiopathic hypersomnia (IH) using various MR methods with instruments selected at $P<5 \times 10^{-8}$. The estimated effects, expressed as odds ratios, are shown as per unit increase in exposure, along with the corresponding 95% confidence intervals (CIs).

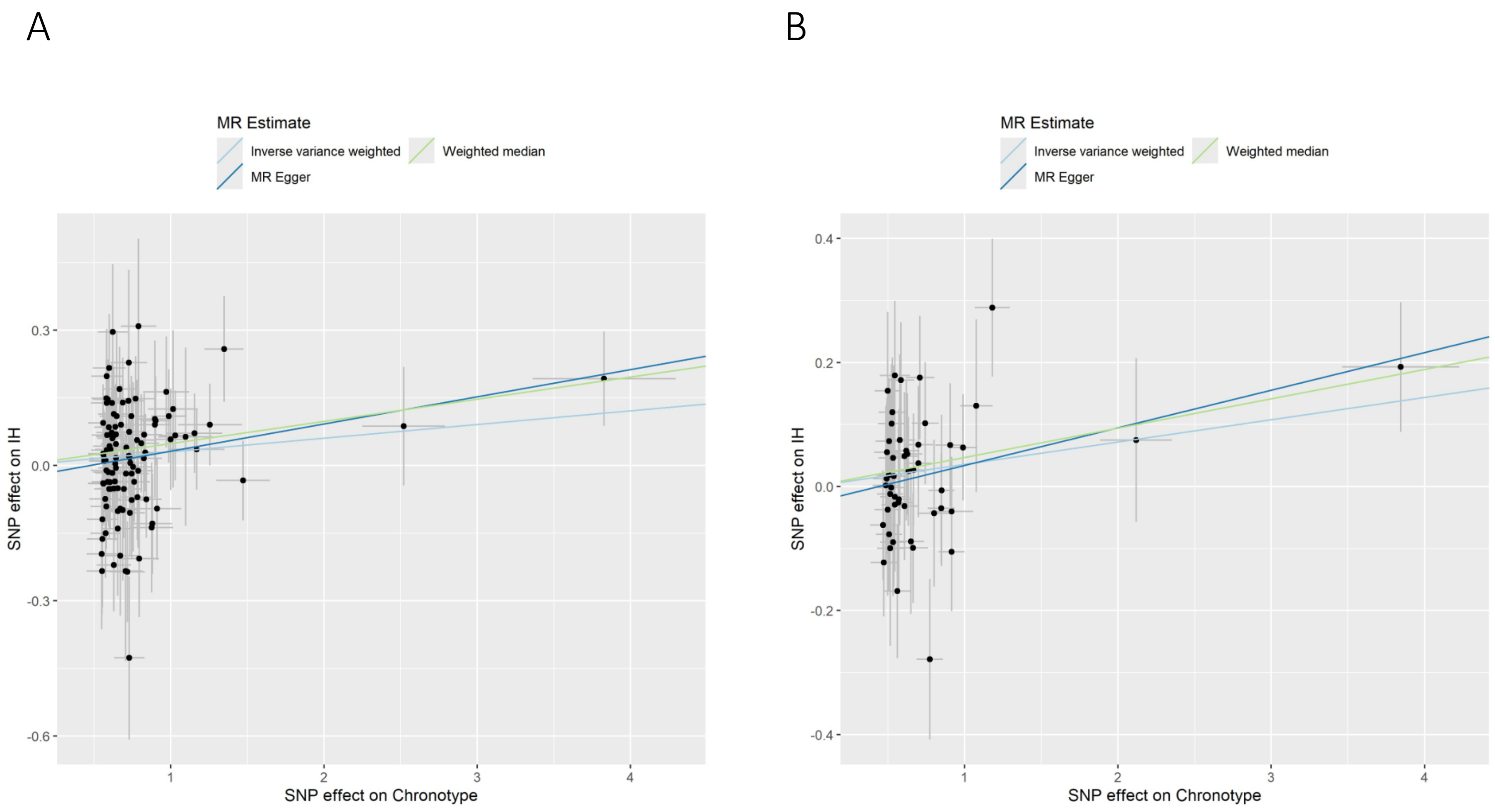


Supplementary Figure S3. MR scatter plots of IH risk against chronotype exposure with genetic instruments selected at $P<5 \times 10^{-8}$. Plots show associations of genetic variants linked to chronotype with IH risk (log odds), plotted against the effects on eveningness (exposure), standardized in standard deviation units. Lines represent the slopes obtained using three different MR methods. Panel (A) shows chronotype results from the UK Biobank, whereas panel (B) presents results from 23andMe. Effect sizes and standard errors (SEs) for chronotype were standardized. Error bars indicate the SEs of the effect estimates.

Supplementary Table S1. Demographic and sleep-related characteristics of idiopathic hypersomnia (IH) patients with objectively confirmed long sleep duration (n = 57) (660 min on 24-h PSG).

| Variable | N | Mean | SD (or SE*) |
|---|---|---|---|
| Age, years | 57 | 22.16 | 7.60 |
| Sex, % female | 57 | 56.1% | 6.6%* |
| BMI | 57 | 20.50 | 3.05 |
| HLA-DQB1*06:02 positivity, % | 57 | 12.3% | 4.4%* |
| Sleep latency (MSLT), min | 57 | 11.86 | 4.05 |
| SOREMP, number | 57 | 0.32 | 0.76 |
| Sleep efficiency, % | 57 | 81.9% | 13.2% |
| Apnea hypopnea index, events/h | 57 | 2.71 | 3.36 |
| Sleep latency (PSG), min | 57 | 36.01 | 59.68 |
| PLMI, events/h | 57 | 1.13 | 2.27 |
| Arousal index, events/h | 57 | 10.96 | 3.87 |
| WASO, % | 57 | 14.0% | 10.7% |
| Total sleep time, min | 57 | 439.57 | 70.13 |
| Stage N1 sleep, min | 57 | 41.86 | 20.55 |
| Stage N2 sleep, min | 57 | 230.33 | 53.99 |
| Stage N3 sleep, min | 57 | 75.90 | 31.16 |
| REM sleep, min | 57 | 91.49 | 29.75 |
| Early morning N3 sleep, % | 56 | 69.6% | 6.1%* |
| JESS in unmedicated conditions | 38 | 15.05 | 4.96 |
| MSFsc | 57 | 5:04 AM | 2 h 16 min |

Data are presented as the mean and standard deviation (SD),

except where indicated by an asterisk (*), which denotes standard error of the mean (SE).

MSLT: multiple sleep latency test; BMI: body mass index; SOREMP: sleep onset rapid eye movement period;

PSG: polysomnography; PLMI: periodic limb movement; WASO: wake after sleep onset;

REM: rapid eye movement; Early morning N3 sleep: N3 within 2 h prior to the scheduled wake time;

JESS: Japanese version of the Epworth Sleepiness Scale; MSFsc: sleep-corrected mid-sleep on free days.

Supplementary Table S2. Demographic and sleep-related characteristics of patients with idiopathic hypersomnia (IH) patients diagnosed using MSLT (n =246).

| Variable | N | Mean | SD (or SE*) |
|---|---|---|---|
| Age, years | 246 | 26.51 | 9.07 |
| Sex, % female | 246 | 56.3% | 3.1%* |
| BMI | 215 | 21.43 | 3.14 |
| HLA-DQB1*06:02 positivity, % | 244 | 10.7% | 1.9%* |
| Sleep latency (MSLT), min | 189 | 4.32 | 2.01 |
| SOREMP, number | 189 | 0.27 | 0.45 |
| Sleep efficiency, % | 190 | 90.5% | 9.7% |
| Apnea hypopnea index, events/h | 193 | 2.24 | 2.67 |
| Sleep latency (PSG), min | 193 | 10.58 | 18.40 |
| PLMI, events/h | 191 | 0.76 | 1.78 |
| Arousal index, events/h | 186 | 11.00 | 6.09 |
| WASO, % | 183 | 8.0% | 9.7% |
| Total sleep time, min | 186 | 501.32 | 75.26 |
| Stage N1 sleep, min | 147 | 45.13 | 24.65 |
| Stage N2 sleep, min | 147 | 270.26 | 51.74 |
| Stage N3 sleep, min | 147 | 71.01 | 38.82 |
| REM sleep, min | 147 | 112.16 | 35.10 |
| Early morning N3 sleep, % | 146 | 41.4% | 4.1%* |
| JESS in unmedicated conditions | 145 | 16.07 | 4.64 |
| MSFsc | 147 | 4:16 AM | 1 h 31 min |

Data are presented as the mean and standard deviation (SD),

except where indicated by an asterisk (*), which denotes standard error of the mean (SE).

MSLT: multiple sleep latency test; BMI: body mass index; SOREMP: sleep onset rapid eye movement period;

PSG: polysomnography; PLMI: periodic limb movement; WASO: wake after sleep onset;

REM: rapid eye movement; Early morning N3 sleep: N3 within 2 h prior to the scheduled wake time;

JESS: Japanese version of the Epworth Sleepiness Scale; MSFsc: sleep-corrected mid-sleep on free days.

Supplementary Table S3. Definitions of sleep traits evaluated in the PRS analysis.

| Sleep Trait | Consortium | Question Stem | Response | Corresponding PGS catalog number |
|---|---|---|---|---|
| Chronotype (morningness/eveningness) | UK Biobank | Do you consider yourself to be? | SELECT one from<br>1 : Definitely a 'morning' person<br>2 : More a 'morning' than 'evening' person<br>3 : More an 'evening' than a 'morning' person<br>4 : Definitely an 'evening' person<br>Participants who answered “do not know” or “prefer not to answer” were treated as missing and excluded. | PGS001055<br>PGS002209<br>PGS002684 |
| Chronotype (morningness/eveningness) | UK Biobank | On an average day, how easy do you find getting up in the morning? | 1 : Not at all easy<br>2 : Not very easy<br>3 : Fairly easy<br>4 : Very easy<br>Participants who answered “do not know” or “prefer not to answer” were treated as missing and excluded. The variables were transformed so that eveningness and difficulty waking up in the morning were coded in the positive direction in the analyses. | PGS001001 |
| Sleep duration | UK Biobank | About how many hours sleep do you get in every 24 hours?<br>(please include naps) | Enter INTEGER | PGS001150<br>PGS002196 |
| Sleeplessness/insomnia | UK biobank | Do you have trouble falling asleep at night or do you wake up in the middle of the night? | SELECT one of 4 from<br>1 : Never/rarely<br>2 : Sometimes<br>3 : Usually<br>Participants who answered “prefer not to answer” were treated as missing and excluded. | PGS001932<br>PGS003328 |
| Daytime napping and dozing | UK Biobank | Do you have a nap during the day? | SELECT one from<br>1 : Never/rarely<br>2 : Sometimes<br>3 : Usually<br>Participants who answered “prefer not to answer” were treated as missing and excluded. | PGS001000 |
| Daytime napping and dozing | UK Biobank | How likely are you to doze off or fall asleep during the daytime when you don't mean to? (e.g., when working, reading or driving) | SELECT one from<br>1 : Never/rarely<br>2 : Sometimes<br>3 : Usually<br>Participants who answered “prefer not to answer” were treated as missing and excluded. | PGS000926<br>PGS002212 |

Supplementary Table S4. Definitions of chronotypes evaluated in MR analyses.

| Sleep Trait | Consortium | Question Stem | Response | Original source |
|---|---|---|---|---|
| Chronotype (morningness/eveningness) | UK Biobank | Do you consider yourself to be? | SELECT one from<br>1 : Definitely a 'morning' person<br>2 : More a 'morning' than 'evening' person<br>3 : More an 'evening' than a 'morning' person<br>4 : Definitely an 'evening' person<br>Participants who answered “do not know” or “prefer not to answer” were treated as missing and excluded. | Exposure data obtained from ukb-b-4956 in the IEU OpenGWAS project (https://doi.org/10.1101/2020.08.10.244293; https://doi.org/10.7554/eLife.34408) |
| Chronotype (morningness/eveningness) | 23andMe | Are you naturally a night person or a morning person? | SELECT one from<br>0 : night owl or night person<br>1 : early bird or morning person<br>Participants who answered “neither”, “it depends” or “I’m not sure” were treated as missing and excluded. The variables were transformed so that eveningness and difficulty waking up in the morning were coded in the positive direction in the analyses. | Exposure data sourced from the original study (https://doi.org/10.1038/s41467-018-08259-7) |

Supplementary Table S5. Summary information for the chronotype genetic instrument (ukb-b-4956, $P < 5 \times 10^{-8}$) used for MR analyses.

| Lead Variant | Phenotype | chr | SNP | effect_allele | other_allele | beta | se | pval | samplesize | eaf |
|---|---|---|---|---|---|---|---|---|---|---|
| rs10058356 | Chronotype | 5 | 35220404 | T | C | 0.627877387 | 0.10644868 | 3.70E-09 | 413343 | 0.697815 |
| rs10118767 | Chronotype | 9 | 126523296 | T | C | 0.669497811 | 0.122387918 | 4.50E-08 | 413343 | 0.197744 |
| rs10149448 | Chronotype | 14 | 84253271 | G | A | 0.553432287 | 0.09985163 | 3.00E-08 | 413343 | 0.396431 |
| rs10175975 | Chronotype | 2 | 59429807 | T | C | -0.880801872 | 0.127194662 | 4.40E-12 | 413343 | 0.181726 |
| rs10280205 | Chronotype | 7 | 24082063 | C | T | 0.638226172 | 0.105593552 | 1.50E-09 | 413343 | 0.309279 |
| rs10402849 | Chronotype | 19 | 2695661 | T | C | -0.727204628 | 0.121765265 | 2.30E-09 | 413343 | 0.201704 |
| rs10460095 | Chronotype | 18 | 22624708 | A | G | -0.642458676 | 0.098869957 | 8.10E-11 | 413343 | 0.574433 |
| rs10737452 | Chronotype | 1 | 21199764 | T | C | 0.687937725 | 0.100564397 | 7.90E-12 | 413343 | 0.624486 |
| rs10742179 | Chronotype | 11 | 27650524 | G | A | 0.618377021 | 0.111112104 | 2.60E-08 | 413343 | 0.739373 |
| rs10954933 | Chronotype | 8 | 33667730 | G | A | -0.742126242 | 0.098849346 | 6.00E-14 | 413343 | 0.42758 |
| rs10988239 | Chronotype | 9 | 131943440 | T | C | 0.617619676 | 0.099164747 | 4.70E-10 | 413343 | 0.512284 |
| rs11183201 | Chronotype | 12 | 46170982 | C | T | -0.619062466 | 0.097785228 | 2.40E-10 | 413343 | 0.508456 |
| rs1135946 | Chronotype | 4 | 83284422 | C | T | 0.827816479 | 0.115273668 | 6.90E-13 | 413343 | 0.232423 |
| rs11587758 | Chronotype | 1 | 150327106 | A | G | -0.900943415 | 0.099364628 | 1.20E-19 | 413343 | 0.396163 |
| rs116131939 | Chronotype | 2 | 50260406 | T | C | -1.095931004 | 0.178337022 | 8.00E-10 | 413343 | 0.083611 |
| rs11714441 | Chronotype | 3 | 123158946 | T | C | 0.565453941 | 0.099487337 | 1.30E-08 | 413343 | 0.400637 |
| rs12117333 | Chronotype | 1 | 179327958 | A | G | 1.154869704 | 0.182263233 | 2.40E-10 | 413343 | 0.077242 |
| rs12249410 | Chronotype | 10 | 64301941 | T | G | 0.910443782 | 0.159098541 | 1.00E-08 | 413343 | 0.109634 |
| rs12377175 | Chronotype | 9 | 8417501 | C | A | 0.744671497 | 0.113203431 | 4.80E-11 | 413343 | 0.22858 |
| rs12432176 | Chronotype | 14 | 101021218 | A | C | -0.579038219 | 0.100941152 | 9.70E-09 | 413343 | 0.379609 |
| rs12525312 | Chronotype | 6 | 165236239 | C | T | 0.58915213 | 0.098036878 | 1.90E-09 | 413343 | 0.550602 |
| rs12713014 | Chronotype | 2 | 48986474 | G | A | 1.256430633 | 0.209077562 | 1.90E-09 | 413343 | 0.058725 |
| rs12811046 | Chronotype | 12 | 24066937 | G | A | 0.621056489 | 0.098093918 | 2.40E-10 | 413343 | 0.445344 |
| rs12965577 | Chronotype | 18 | 31675680 | G | A | 0.760892102 | 0.10349839 | 2.00E-13 | 413343 | 0.334996 |
| rs12969848 | Chronotype | 18 | 38152835 | T | C | -0.787154719 | 0.098000928 | 9.60E-16 | 413343 | 0.529314 |
| rs12971913 | Chronotype | 19 | 18441704 | A | G | 0.607026432 | 0.098301469 | 6.60E-10 | 413343 | 0.448148 |
| rs13011556 | Chronotype | 2 | 4651923 | G | C | -0.808801366 | 0.114702303 | 1.80E-12 | 413343 | 0.239067 |
| rs13059636 | Chronotype | 3 | 24935790 | G | A | -0.642108763 | 0.09810734 | 6.00E-11 | 413343 | 0.469784 |
| rs13316611 | Chronotype | 3 | 63564385 | T | G | -0.648033311 | 0.111829186 | 6.80E-09 | 413343 | 0.255868 |
| rs139911 | Chronotype | 22 | 40704052 | T | C | 0.84124737 | 0.099059294 | 2.00E-17 | 413343 | 0.575785 |
| rs1421085 | Chronotype | 16 | 53800954 | C | T | -0.999259265 | 0.099393388 | 8.90E-24 | 413343 | 0.403565 |
| rs1439319 | Chronotype | 15 | 48008970 | C | G | 0.583117337 | 0.102752549 | 1.40E-08 | 413343 | 0.645445 |
| rs147762489 | Chronotype | 5 | 87603572 | T | C | 0.7807077 | 0.113143515 | 5.20E-12 | 413343 | 0.249106 |
| rs17161045 | Chronotype | 7 | 32261458 | C | T | 0.735420383 | 0.101671175 | 4.70E-13 | 413343 | 0.369932 |
| rs17448682 | Chronotype | 1 | 15966713 | T | C | -0.790198479 | 0.115529152 | 7.90E-12 | 413343 | 0.232109 |
| rs17517 | Chronotype | 13 | 109790422 | A | G | 0.55802429 | 0.098231966 | 1.30E-08 | 413343 | 0.511747 |
| rs17575798 | Chronotype | 1 | 110086451 | A | G | 0.76978851 | 0.123110271 | 4.00E-10 | 413343 | 0.193371 |
| rs1800828 | Chronotype | 3 | 113891549 | G | C | 0.628639526 | 0.112046802 | 2.00E-08 | 413343 | 0.253098 |
| rs1874493 | Chronotype | 19 | 31699729 | G | A | 0.589597909 | 0.104655978 | 1.80E-08 | 413343 | 0.680061 |
| rs1914772 | Chronotype | 11 | 43884417 | A | T | 1.014291126 | 0.16144679 | 3.30E-10 | 413343 | 0.894653 |
| rs197273 | Chronotype | 2 | 161894663 | G | A | 0.579781184 | 0.097808716 | 3.10E-09 | 413343 | 0.529834 |
| rs1976423 | Chronotype | 5 | 104042643 | C | A | -0.670916635 | 0.097652453 | 6.40E-12 | 413343 | 0.495282 |
| rs1983891 | Chronotype | 6 | 41536427 | T | C | 0.618760487 | 0.107438981 | 8.50E-09 | 413343 | 0.276348 |
| rs2072727 | Chronotype | 20 | 43538733 | C | T | 0.558671389 | 0.09842322 | 1.40E-08 | 413343 | 0.564423 |
| rs2077432 | Chronotype | 11 | 66621383 | T | C | -0.642209423 | 0.110233488 | 5.70E-09 | 413343 | 0.269742 |
| rs2239626 | Chronotype | 3 | 185998884 | C | T | -0.666765617 | 0.106128966 | 3.30E-10 | 413343 | 0.305676 |
| rs2291589 | Chronotype | 9 | 37079661 | G | T | 0.728148912 | 0.100564397 | 4.50E-13 | 413343 | 0.377049 |
| rs2364972 | Chronotype | 17 | 65475332 | G | A | -0.613650805 | 0.097953953 | 3.70E-10 | 413343 | 0.462953 |
| rs2518022 | Chronotype | 17 | 8057367 | C | T | 1.471665218 | 0.173892174 | 2.60E-17 | 413343 | 0.914438 |
| rs2701524 | Chronotype | 15 | 37383688 | C | T | 0.550882239 | 0.099648393 | 3.20E-08 | 413343 | 0.413565 |
| rs2712056 | Chronotype | 2 | 23959131 | T | C | -0.753999304 | 0.125377034 | 1.80E-09 | 413343 | 0.185573 |
| rs2762088 | Chronotype | 13 | 94065186 | G | T | -0.69345484 | 0.114828847 | 1.60E-09 | 413343 | 0.761336 |
| rs2850298 | Chronotype | 2 | 44731590 | G | A | -0.895991913 | 0.10612561 | 3.10E-17 | 413343 | 0.698902 |
| rs286808 | Chronotype | 5 | 107459376 | C | T | 0.561422756 | 0.097787146 | 9.40E-09 | 413343 | 0.524555 |
| rs2881955 | Chronotype | 6 | 72479263 | T | C | -0.653459352 | 0.108973803 | 2.00E-09 | 413343 | 0.278459 |
| rs2893787 | Chronotype | 10 | 60568697 | A | G | 0.710010978 | 0.111544462 | 1.90E-10 | 413343 | 0.743579 |
| rs2971970 | Chronotype | 7 | 133643778 | G | T | 0.726504803 | 0.118246007 | 8.00E-10 | 413343 | 0.782046 |
| rs3100052 | Chronotype | 8 | 101967139 | G | A | 0.584943593 | 0.100163196 | 5.20E-09 | 413343 | 0.613151 |
| rs3168135 | Chronotype | 11 | 58386177 | A | G | 0.717579635 | 0.114063833 | 3.20E-10 | 413343 | 0.24042 |

| | | | | | | | | | | |
|---|---|---|---|---|---|---|---|---|---|---|
| rs34244172 | Chronotype | 3 | 172339648 | T | C | 0.598058125 | 0.10781334 | 2.90E-08 | 413343 | 0.292264 |
| rs35101255 | Chronotype | 6 | 110235958 | G | A | 1.170265858 | 0.180283112 | 8.50E-11 | 413343 | 0.079468 |
| rs3808964 | Chronotype | 10 | 125426627 | T | G | -0.580898028 | 0.101352899 | 1.00E-08 | 413343 | 0.633453 |
| rs4141920 | Chronotype | 11 | 30449319 | A | G | 0.558537176 | 0.098035919 | 1.20E-08 | 413343 | 0.455163 |
| rs4237555 | Chronotype | 11 | 92725803 | T | C | -0.560909871 | 0.097695114 | 9.40E-09 | 413343 | 0.527732 |
| rs4241964 | Chronotype | 4 | 137053959 | G | T | -0.729831369 | 0.098131306 | 1.00E-13 | 413343 | 0.475791 |
| rs4321976 | Chronotype | 8 | 76614958 | C | T | 0.824715199 | 0.117543785 | 2.30E-12 | 413343 | 0.221209 |
| rs4484214 | Chronotype | 3 | 182257586 | G | A | 0.653607946 | 0.104980965 | 4.80E-10 | 413343 | 0.314742 |
| rs4518438 | Chronotype | 5 | 88157552 | C | T | -0.731652832 | 0.097534538 | 6.30E-14 | 413343 | 0.510399 |
| rs4549082 | Chronotype | 2 | 198944654 | C | T | -0.733790654 | 0.097576239 | 5.50E-14 | 413343 | 0.484139 |
| rs4671379 | Chronotype | 2 | 60495038 | C | T | -0.593499674 | 0.098971096 | 2.00E-09 | 413343 | 0.58559 |
| rs4784655 | Chronotype | 16 | 56364030 | C | G | 0.783205021 | 0.10470487 | 7.40E-14 | 413343 | 0.321967 |
| rs486416 | Chronotype | 6 | 31856070 | A | G | 0.554990117 | 0.100782493 | 3.70E-08 | 413343 | 0.636771 |
| rs4886947 | Chronotype | 15 | 78166330 | A | G | 0.571915341 | 0.10369971 | 3.50E-08 | 413343 | 0.660074 |
| rs4936291 | Chronotype | 11 | 114011665 | G | A | -0.653291586 | 0.102767888 | 2.10E-10 | 413343 | 0.389142 |
| rs56372114 | Chronotype | 1 | 79867495 | T | C | 0.581410914 | 0.100103759 | 6.30E-09 | 413343 | 0.384232 |
| rs6131942 | Chronotype | 20 | 17348608 | G | A | -0.685857423 | 0.099100516 | 4.50E-12 | 413343 | 0.579505 |
| rs62182115 | Chronotype | 2 | 240266397 | T | C | 0.636802555 | 0.102663873 | 5.50E-10 | 413343 | 0.341565 |
| rs62198772 | Chronotype | 2 | 186189829 | A | T | 0.566460539 | 0.099544857 | 1.30E-08 | 413343 | 0.410702 |
| rs6442446 | Chronotype | 3 | 14394752 | G | A | 0.624109835 | 0.107875173 | 7.20E-09 | 413343 | 0.708837 |
| rs6504758 | Chronotype | 17 | 50208366 | G | A | -0.597713005 | 0.098266478 | 1.20E-09 | 413343 | 0.535685 |
| rs6658041 | Chronotype | 1 | 79387676 | A | G | -0.575328187 | 0.099553485 | 7.50E-09 | 413343 | 0.599474 |
| rs66710942 | Chronotype | 3 | 77215327 | T | C | -0.637837913 | 0.099681946 | 1.60E-10 | 413343 | 0.592333 |
| rs6718119 | Chronotype | 2 | 76341929 | G | A | -0.574537288 | 0.100561042 | 1.10E-08 | 413343 | 0.376746 |
| rs67988891 | Chronotype | 5 | 152204741 | G | C | -0.897180657 | 0.10481943 | 1.10E-17 | 413343 | 0.318639 |
| rs6967481 | Chronotype | 7 | 50642701 | T | C | -0.744149025 | 0.097932383 | 3.00E-14 | 413343 | 0.497073 |
| rs698015 | Chronotype | 14 | 57269825 | T | C | -0.62958381 | 0.103571249 | 1.20E-09 | 413343 | 0.646935 |
| rs72632979 | Chronotype | 11 | 16615883 | G | A | 0.793175133 | 0.129553936 | 9.20E-10 | 413343 | 0.172073 |
| rs72720396 | Chronotype | 1 | 91191582 | G | A | -0.988292141 | 0.11573143 | 1.30E-17 | 413343 | 0.229643 |
| rs7304278 | Chronotype | 12 | 106989915 | G | A | -0.705735334 | 0.109433003 | 1.10E-10 | 413343 | 0.723741 |
| rs74097630 | Chronotype | 12 | 63514274 | T | G | 0.874560965 | 0.140516265 | 4.80E-10 | 413343 | 0.140832 |
| rs74357745 | Chronotype | 11 | 122811822 | G | A | 0.970077514 | 0.149860849 | 9.60E-11 | 413343 | 0.12053 |
| rs7547493 | Chronotype | 1 | 77775233 | G | A | -1.347580464 | 0.127376329 | 3.70E-26 | 413343 | 0.177307 |
| rs76223855 | Chronotype | 6 | 153136141 | C | T | -3.827827961 | 0.46758435 | 2.70E-16 | 413343 | 0.010981 |
| rs7626349 | Chronotype | 3 | 138130484 | C | T | -0.601006018 | 0.106976905 | 1.90E-08 | 413343 | 0.705763 |
| rs7783012 | Chronotype | 7 | 114116881 | A | G | -0.581027448 | 0.099455222 | 5.20E-09 | 413343 | 0.590654 |
| rs78095690 | Chronotype | 20 | 16239683 | C | T | -0.55063778 | 0.098634605 | 2.40E-08 | 413343 | 0.437331 |
| rs7959983 | Chronotype | 12 | 90452978 | C | T | -0.67487592 | 0.09893994 | 9.00E-12 | 413343 | 0.405084 |
| rs812925 | Chronotype | 2 | 61680993 | G | C | -0.724635407 | 0.10185476 | 1.10E-12 | 413343 | 0.351718 |
| rs9291813 | Chronotype | 5 | 63842133 | C | T | -0.641624638 | 0.11420236 | 1.90E-08 | 413343 | 0.760663 |
| rs9348050 | Chronotype | 6 | 166263488 | C | T | 0.597094667 | 0.097601644 | 9.50E-10 | 413343 | 0.510601 |
| rs9395520 | Chronotype | 6 | 13183523 | T | C | -0.832667322 | 0.106044603 | 4.10E-15 | 413343 | 0.304164 |
| rs9476310 | Chronotype | 6 | 57767576 | T | C | -0.603997052 | 0.097752154 | 6.50E-10 | 413343 | 0.51056 |
| rs9573971 | Chronotype | 13 | 77568732 | G | A | 2.519739685 | 0.269811362 | 9.70E-21 | 413343 | 0.03387 |
| rs9795439 | Chronotype | 11 | 1483543 | G | A | 0.708242242 | 0.122747417 | 7.90E-09 | 413343 | 0.803776 |
| rs9932577 | Chronotype | 16 | 24512509 | A | C | 0.60641768 | 0.09848074 | 7.40E-10 | 413343 | 0.505674 |
| rs9964420 | Chronotype | 18 | 56824041 | A | C | 1.029984465 | 0.106598711 | 4.40E-22 | 413343 | 0.302896 |

se: standard error; eaf: effect allele frequency.

When genetic variants with $P < 6\times10^{-9}$ were examined, they were selected by sorting *P* values in ascending order.

Positive beta value indicates a genetic tendency toward eveningness.

Supplementary Table S6. Summary information for the chronotype genetic instrument (23andMe, $P < 5\times10^{-8}$) used for MR analyses.

| Lead Variant | Phenotype | chr | SNP | effect_allele | other_allele | beta | se | pval | samplesize | eaf |
|---|---|---|---|---|---|---|---|---|---|---|
| rs10917513 | Chronotype | 1 | 20006887 | T | C | 0.487246252 | 0.087819541 | 2.88E-08 | 248094 | 0.6476 |
| rs10916892 | Chronotype | 1 | 21201325 | T | C | 0.515517024 | 0.086385569 | 2.40E-09 | 248094 | 0.6203 |
| rs12040629 | Chronotype | 1 | 77705365 | A | G | -1.180131154 | 0.11459978 | 7.07E-25 | 248094 | 0.16 |
| rs9436119 | Chronotype | 1 | 150467753 | A | G | -0.742593836 | 0.089411035 | 9.87E-17 | 248094 | 0.3842 |
| rs146820337 | Chronotype | 1 | 190095126 | C | CTAACA | 0.56222255 | 0.08410035 | 2.30E-11 | 248094 | 0.4751 |
| rs359248 | Chronotype | 2 | 60477461 | T | G | 0.533383337 | 0.084778945 | 3.13E-10 | 248094 | 0.4564 |
| rs10520176 | Chronotype | 2 | 77217310 | T | C | -0.545577962 | 0.086664417 | 3.06E-10 | 248094 | 0.4926 |
| rs6433478 | Chronotype | 2 | 175241482 | T | C | 0.498674471 | 0.086065291 | 6.85E-09 | 248094 | 0.4601 |
| rs1064213 | Chronotype | 2 | 198950240 | A | G | -0.914550209 | 0.084307364 | 1.99E-27 | 248094 | 0.4826 |
| rs7429614 | Chronotype | 3 | 77205438 | T | G | -0.64989687 | 0.085961501 | 4.00E-14 | 248094 | 0.416 |
| rs3850174 | Chronotype | 3 | 172364093 | A | T | 0.545620383 | 0.097952081 | 2.53E-08 | 248094 | 0.2575 |
| rs9836621 | Chronotype | 3 | 182096311 | T | C | 0.570480506 | 0.084156911 | 1.21E-11 | 248094 | 0.5217 |
| rs4698678 | Chronotype | 4 | 18260776 | C | G | -0.534069143 | 0.09553168 | 2.26E-08 | 248094 | 0.2786 |
| rs17455138 | Chronotype | 4 | 130903511 | T | C | -0.578421718 | 0.100002995 | 7.25E-09 | 248094 | 0.7655 |
| rs67169439 | Chronotype | 5 | 59027048 | T | TA | 0.499268365 | 0.086375106 | 7.45E-09 | 248094 | 0.6014 |
| rs4269995 | Chronotype | 5 | 87701223 | T | C | 0.663856209 | 0.097727956 | 1.09E-11 | 248094 | 0.251 |
| rs1559253 | Chronotype | 5 | 106657015 | A | G | -0.508222025 | 0.088810778 | 1.05E-08 | 248094 | 0.3588 |
| rs34125199 | Chronotype | 6 | 12155114 | T | TTC | -0.524511691 | 0.088179271 | 2.71E-09 | 248094 | 0.3497 |
| rs9381812 | Chronotype | 6 | 13183998 | A | G | 0.989930921 | 0.092488538 | 9.63E-27 | 248094 | 0.7051 |
| rs12195792 | Chronotype | 6 | 98705295 | A | T | -0.666456616 | 0.094194004 | 1.49E-12 | 248094 | 0.2717 |
| rs9479402 | Chronotype | 6 | 153135339 | T | C | 3.844417737 | 0.382354943 | 6.75E-24 | 248094 | 0.9883 |
| rs10237162 | Chronotype | 7 | 24085405 | T | C | -0.699242397 | 0.093806559 | 8.94E-14 | 248094 | 0.7232 |
| rs10951325 | Chronotype | 7 | 32265545 | T | C | -0.522567395 | 0.087635999 | 2.47E-09 | 248094 | 0.6319 |
| rs6967481 | Chronotype | 7 | 50642701 | T | C | -0.507151601 | 0.08509894 | 2.52E-09 | 248094 | 0.5011 |
| rs10254050 | Chronotype | 7 | 96468077 | C | G | 1.07614313 | 0.106172282 | 3.64E-24 | 248094 | 0.1905 |
| rs6958557 | Chronotype | 7 | 133585794 | T | G | -0.494651545 | 0.086075048 | 9.06E-09 | 248094 | 0.6067 |
| rs6993892 | Chronotype | 8 | 33729200 | T | C | 0.528220701 | 0.087310206 | 1.45E-09 | 248094 | 0.6162 |
| rs34054660 | Chronotype | 8 | 65015659 | A | G | -0.469030672 | 0.085378919 | 3.93E-08 | 248094 | 0.5747 |
| rs7006885 | Chronotype | 8 | 93283578 | A | G | -0.708781467 | 0.093995474 | 4.66E-14 | 248094 | 0.2878 |
| rs6477309 | Chronotype | 9 | 8450638 | T | C | -0.491824892 | 0.089416409 | 3.78E-08 | 248094 | 0.6661 |
| rs9416744 | Chronotype | 10 | 60567937 | A | C | -0.626802875 | 0.096545117 | 8.45E-11 | 248094 | 0.2595 |
| rs60521023 | Chronotype | 11 | 13314102 | A | AT | -0.606750465 | 0.092920808 | 6.58E-11 | 248094 | 0.2965 |
| rs10742179 | Chronotype | 11 | 27650524 | A | G | -0.543864153 | 0.095306141 | 1.15E-08 | 248094 | 0.2627 |
| rs10838687 | Chronotype | 11 | 47312892 | T | G | -0.634322705 | 0.104145972 | 1.12E-09 | 248094 | 0.7918 |
| rs662094 | Chronotype | 11 | 66342691 | A | G | -0.506221167 | 0.084323202 | 1.93E-09 | 248094 | 0.4944 |
| rs1278402 | Chronotype | 11 | 82972097 | A | G | -0.537540595 | 0.096012027 | 2.15E-08 | 248094 | 0.736 |
| rs1508608 | Chronotype | 11 | 92893825 | A | G | -0.568737003 | 0.09018734 | 2.85E-10 | 248094 | 0.3199 |
| rs13377754 | Chronotype | 12 | 34051765 | T | C | -0.772622252 | 0.086935911 | 6.15E-19 | 248094 | 0.6104 |
| rs1843888 | Chronotype | 12 | 38737310 | A | G | -0.849443864 | 0.084926145 | 1.46E-23 | 248094 | 0.5433 |
| rs10877962 | Chronotype | 12 | 63520912 | T | C | -0.698560833 | 0.086778812 | 8.23E-16 | 248094 | 0.4083 |
| rs711098 | Chronotype | 12 | 77976559 | A | C | -0.47281604 | 0.086676436 | 4.89E-08 | 248094 | 0.3979 |
| rs9573980 | Chronotype | 13 | 77590741 | A | G | -2.118476719 | 0.234323876 | 1.36E-19 | 248094 | 0.9659 |
| rs1873958 | Chronotype | 15 | 101147726 | A | G | -0.585390075 | 0.085929261 | 9.56E-12 | 248094 | 0.4088 |
| rs1421085 | Chronotype | 16 | 53800954 | T | C | 0.620411443 | 0.085549169 | 4.08E-13 | 248094 | 0.5935 |
| rs2550298 | Chronotype | 16 | 56367969 | T | C | 0.606985195 | 0.088943132 | 8.77E-12 | 248094 | 0.3811 |
| rs1061032 | Chronotype | 17 | 8064083 | T | G | -0.914342346 | 0.140896985 | 8.61E-11 | 248094 | 0.094 |
| rs4419127 | Chronotype | 18 | 31663654 | A | G | -0.847806414 | 0.088670223 | 1.13E-21 | 248094 | 0.6622 |
| rs12969848 | Chronotype | 18 | 38152835 | T | C | -0.513893713 | 0.084822497 | 1.37E-09 | 248094 | 0.5313 |
| rs4800998 | Chronotype | 18 | 53429655 | A | T | -0.801798005 | 0.110783304 | 4.56E-13 | 248094 | 0.1827 |
| rs9964420 | Chronotype | 18 | 56824041 | A | C | 0.905264251 | 0.094916575 | 1.42E-21 | 248094 | 0.2983 |

se: standard error; eaf: effect allele frequency.

When genetic variants with $P < 6\times10^{-9}$ were examined, they were selected by sorting the $P$ values in ascending order.
Positive beta value indicates a genetic tendency toward eveningness.

Supplementary Table S7. Associations between PRS percentiles for sleep-related traits and risk of IH.

| Top PRS for chronotype | IH patients (n=303) | Controls (n=2918) | OR | 95% CI | | *P* | Bottom PRS for chronotype | IH patients (n=303) | Controls (n=2918) | OR | 95% CI | | *P* | *P* (top vs. bottom) |
|---|---|---|---|---|---|---|---|---|---|---|---|---|---|---|
| 0.5% | 14 | 41 | 3.40 | 1.83 | 6.31 | 3.9E-05 | 0.5% | 3 | 53 | 0.45 | 0.14 | 1.44 | 0.17 | 7.3E-03 |
| 1.0% | 21 | 85 | 2.48 | 1.52 | 4.06 | 1.9E-04 | 1.0% | 7 | 101 | 0.55 | 0.25 | 1.18 | 0.12 | 5.8E-03 |
| 2.5% | 37 | 219 | 1.71 | 1.18 | 2.48 | 3.9E-03 | 2.5% | 14 | 241 | 0.44 | 0.26 | 0.77 | 2.9E-03 | 1.1E-03 |
| 5.0% | 67 | 396 | 1.81 | 1.35 | 2.42 | 5.5E-05 | 5.0% | 33 | 434 | 0.57 | 0.39 | 0.83 | 2.6E-03 | 3.5E-04 |

| Top PRS for daytime napping | IH patients (n=303) | Controls (n=2918) | OR | 95% CI | | *P* | Bottom PRS for daytime napping | IH patients (n=303) | Controls (n=2918) | OR | 95% CI | | *P* | *P* (top vs. bottom) |
|---|---|---|---|---|---|---|---|---|---|---|---|---|---|---|
| 0.5% | 13 | 34 | 3.80 | 1.98 | 7.29 | 1.6E-05 | 0.5% | 2 | 41 | 0.47 | 0.11 | 1.94 | 0.28 | 9.5E-03 |
| 1.0% | 15 | 79 | 1.87 | 1.06 | 3.29 | 0.027 | 1.0% | 4 | 84 | 0.45 | 0.16 | 1.24 | 0.11 | 0.018 |
| 2.5% | 28 | 186 | 1.50 | 0.99 | 2.27 | 0.057 | 2.5% | 13 | 201 | 0.61 | 0.34 | 1.08 | 0.084 | 0.016 |
| 5.0% | 46 | 357 | 1.28 | 0.92 | 1.79 | 0.14 | 5.0% | 33 | 374 | 0.83 | 0.57 | 1.21 | 0.34 | 0.11 |

| Top PRS for sleep duration | IH patients (n=303) | Controls (n=2918) | OR | 95% CI | | *P* | Bottom PRS for sleep duration | IH patients (n=303) | Controls (n=2918) | OR | 95% CI | | *P* | *P* (top vs. bottom) |
|---|---|---|---|---|---|---|---|---|---|---|---|---|---|---|
| 0.5% | 5 | 26 | 1.87 | 0.71 | 4.90 | 0.20 | 0.5% | 2 | 27 | 0.71 | 0.17 | 3.01 | 0.64 | 0.28 |
| 1.0% | 6 | 55 | 1.05 | 0.45 | 2.46 | 0.91 | 1.0% | 4 | 56 | 0.68 | 0.25 | 1.90 | 0.46 | 0.53 |
| 2.5% | 13 | 135 | 0.92 | 0.52 | 1.65 | 0.79 | 2.5% | 15 | 129 | 1.13 | 0.65 | 1.95 | 0.67 | 0.64 |
| 5.0% | 27 | 256 | 1.02 | 0.67 | 1.54 | 0.94 | 5.0% | 23 | 255 | 0.86 | 0.55 | 1.34 | 0.50 | 0.60 |

| Top PRS for insomnia | IH patients (n=303) | Controls (n=2918) | OR | 95% CI | | *P* | Bottom PRS for insomnia | IH patients (n=303) | Controls (n=2918) | OR | 95% CI | | *P* | *P* (top vs. bottom) |
|---|---|---|---|---|---|---|---|---|---|---|---|---|---|---|
| 0.5% | 4 | 24 | 1.61 | 0.56 | 4.68 | 0.37 | 0.5% | 5 | 26 | 1.87 | 0.71 | 4.90 | 0.20 | 0.84 |
| 1.0% | 4 | 50 | 0.77 | 0.28 | 2.14 | 0.61 | 1.0% | 6 | 53 | 1.09 | 0.47 | 2.56 | 0.84 | 0.61 |
| 2.5% | 13 | 125 | 1.00 | 0.56 | 1.80 | 1.00 | 2.5% | 18 | 124 | 1.42 | 0.86 | 2.37 | 0.17 | 0.39 |
| 5.0% | 24 | 237 | 0.97 | 0.63 | 1.51 | 0.90 | 5.0% | 29 | 235 | 1.21 | 0.81 | 1.81 | 0.36 | 0.50 |

OR: odds ratio; 95% CI: 95% confidence interval for the odds ratio.

Supplementary Table S8. Associations between PRSs for sleep-related traits and IH: comparison of mean adjusted Z-scores and odds ratios.

| | Mean adjusted Z-score of PRS (IH) | Mean adjusted Z-score of PRS (control) | *P* | OR | 95% CI | |
|---|---|---|---|---|---|---|
| Chronotype | 0.234 | -0.024 | 3.0E-05 | 1.18 | 1.09 | 1.28 |
| Daytime napping | 0.134 | -0.014 | 9.8E-03 | 1.13 | 1.03 | 1.25 |
| Sleep duration | 0.031 | -0.003 | 0.52 | 1.03 | 0.93 | 1.14 |
| Insomnia | 0.016 | -0.002 | 0.86 | 1.01 | 0.92 | 1.11 |

OR: odds ratio; 95% CI: 95% confidence interval for the odds ratio.

Supplementary Table S9. Comparison of sleep-related traits in IH patients with extreme PRS values for eveningness and morningness chronotypes (top and bottom 5%).

| Variable | Eve PRS: Mean | Eve PRS: SD (or SE*) | Mor PRS: Mean | Mor PRS: SD (or SE*) | *P* (Eve vs. Mor) | Eve PRS: N | Mor PRS: N | *P* (Eve vs. Others) | *P* (Mor vs. Others) |
|---|---|---|---|---|---|---|---|---|---|
| Mean sleep latency (MSLT), min | 6.37 | 4.55 | 5.07 | 4.25 | 0.31 | 53 | 27 | 0.73 | 0.11 |
| SOREMP (Yes/No) (MSLT), % | 24.5% | 43.4% | 48.1% | 89.3% | 0.14 | 53 | 27 | 0.62 | 0.098 |
| Sleep efficiency (PSG), % | 87.5% | 11.9% | 89.0% | 10.7% | 0.67 | 55 | 27 | 0.66 | 0.89 |
| Apnea hypopnea index (PSG), events/h | 2.73 | 2.96 | 2.05 | 2.61 | 0.32 | 55 | 27 | 0.47 | 0.37 |
| Sleep latency (PSG), min | 14.27 | 18.24 | 18.17 | 37.58 | 0.51 | 55 | 27 | 0.50 | 0.78 |
| PLMI (PSG), events/h | 1.41 | 2.83 | 0.96 | 1.57 | 0.40 | 54 | 27 | 0.13 | 0.91 |
| Arousal index (PSG), events/h | 10.84 | 6.85 | 11.14 | 4.03 | 0.84 | 54 | 27 | 0.82 | 0.98 |
| WASO (PSG), % | 10.6% | 11.5% | 10.2% | 13.3% | 0.93 | 54 | 27 | 0.53 | 0.64 |
| Total sleep time (PSG), min | 491.39 | 91.48 | 478.78 | 87.15 | 0.37 | 54 | 27 | 0.53 | 0.52 |
| Stage N1 sleep (PSG), min | 44.85 | 26.39 | 40.59 | 19.86 | 0.49 | 50 | 23 | 0.86 | 0.44 |
| Stage N2 sleep (PSG), min | 261.47 | 62.98 | 264.59 | 43.74 | 0.97 | 50 | 23 | 0.66 | 0.69 |
| Stage N3 sleep (PSG), min | 78.16 | 39.01 | 80.59 | 41.45 | 0.98 | 50 | 23 | 0.26 | 0.30 |
| REM sleep (PSG), min | 107.88 | 41.47 | 112.30 | 24.32 | 0.72 | 50 | 23 | 0.70 | 0.50 |
| Early morning N3 sleep (PSG), % | 51.1% | 7.4%* | 72.7% | 9.7%* | 0.030 | 47 | 22 | 0.96 | 0.04 |
| JESS in unmedicated conditions | 14.80 | 6.08 | 15.14 | 4.76 | 0.68 | 41 | 21 | 0.38 | 0.74 |
| MSFsc | 4:44 AM | 2 h 3 min | 4:12 AM | 1 h 59 min | 0.34 | 50 | 23 | 0.47 | 0.44 |

Data are presented as the mean and standard deviation (SD), except where indicated by an asterisk (*), which denotes standard error of the mean (SE).

MSLT: multiple sleep latency test; SOREMP: sleep onset rapid eye movement period; PSG: polysomnography; PLMI: periodic limb movement; WASO: wake after sleep onset; REM: rapid eye movement; Early morning N3 sleep: N3 within 2 h prior to the scheduled wake time; JESS: Japanese version of the Epworth Sleepiness Scale; MSFsc: midpoint of sleep-corrected for free days.

Supplementary Table S10. Comparison of sleep-related traits between patients in the top and bottom 2.5% PRS groups for daytime napping (nap).

| Variable | Top NAP PRS: Mean | Top NAP PRS: SD (or SE*) | Bottom NAP PRS: Mean | Bottom NAP PRS: SD (or SE*) | *P* (Top NAP vs. Bottom NAP PRS) | Top NAP PRS: N | Bottom NAP PRS: N | *P* (Top NAP PRS vs. Others) | *P* (Bottom NAP PRS vs. Others) |
|---|---|---|---|---|---|---|---|---|---|
| Mean sleep latency (MSLT), min | 6.03 | 4.75 | 5.98 | 4.02 | 0.37 | 21 | 12 | 0.33 | 0.94 |
| SOREMP (Yes/No) (MSLT), % | 19.0% | 40.2% | 33.3% | 49.2% | 0.37 | 21 | 12 | 0.38 | 0.705 |
| Sleep efficiency (PSG), % | 87.4% | 17.6% | 89.8% | 10.1% | 0.47 | 19 | 13 | 0.45 | 0.71 |
| Apnea hypopnea index (PSG), events/h | 1.63 | 1.25 | 1.90 | 1.50 | 0.73 | 20 | 13 | 0.37 | 0.59 |
| Sleep latency (PSG), min | 31.98 | 96.49 | 12.96 | 14.13 | 0.49 | 20 | 13 | 0.08 | 0.75 |
| PLMI (PSG), events/h | 1.33 | 2.82 | 1.92 | 2.93 | 0.69 | 20 | 12 | 0.35 | 0.09 |
| Arousal index (PSG), events/h | 9.90 | 4.60 | 9.69 | 2.83 | 0.80 | 19 | 13 | 0.52 | 0.42 |
| WASO (PSG), % | 7.7% | 4.6% | 7.9% | 8.1% | 0.77 | 19 | 13 | 0.68 | 0.58 |
| Total sleep time (PSG), min | 471.13 | 98.26 | 499.92 | 73.88 | 0.23 | 19 | 13 | 0.22 | 0.54 |
| Stage N1 sleep (PSG), min | 38.41 | 21.63 | 34.11 | 13.66 | 0.41 | 16 | 9 | 0.41 | 0.20 |
| Stage N2 sleep (PSG), min | 255.16 | 72.40 | 263.06 | 55.81 | 0.32 | 16 | 9 | 0.59 | 0.75 |
| Stage N3 sleep (PSG), min | 62.81 | 36.90 | 76.00 | 35.54 | 0.26 | 16 | 9 | 0.15 | 0.68 |
| REM sleep (PSG), min | 106.97 | 32.81 | 100.83 | 38.66 | 0.72 | 16 | 9 | 0.86 | 0.68 |
| Early morning N3 sleep (PSG), % | 13.3% | 9.1%* | 66.7% | 16.7%* | 0.08 | 15 | 9 | 0.018 | 0.26 |
| JESS in unmedicated conditions | 15.88 | 5.38 | 16.60 | 5.23 | 0.81 | 16 | 10 | 0.86 | 0.76 |
| MSFsc | 4:35 AM | 1 h 55 min | 5:11 AM | 2 h 54 min | 0.95 | 16 | 9 | 0.86 | 0.25 |

Data are presented as the mean and standard deviation (SD), except where indicated by an asterisk (*), which denotes standard error of the mean (SE).

MSLT: multiple sleep latency test; SOREMP: sleep onset rapid eye movement period; PSG: polysomnography; PLMI: periodic limb movement; WASO: wake after sleep onset; REM: rapid eye movement; Early morning N3 sleep: N3 within 2 h prior to the scheduled wake time; JESS: Japanese version of the Epworth Sleepiness Scale; MSFsc: midpoint of sleep-corrected for free days.

Supplementary Table S11. Comparison of sleep-related traits between patients in the top and bottom 5% PRS groups for daytime napping (nap).

| Variable | Top NAP PRS: Mean | Top NAP PRS: SD (or SE*) | Bottom NAP PRS: Mean | Bottom NAP PRS: SD (or SE*) | *P* (Top NAP vs. Bottom NAP PRS) | Top NAP PRS: N | Bottom NAP PRS: N | *P* (Top NAP PRS vs. Others) | *P* (Bottom NAP PRS vs. Others) |
|---|---|---|---|---|---|---|---|---|---|
| Mean sleep latency (MSLT), min | 6.44 | 5.12 | 5.89 | 3.58 | 0.12 | 35 | 30 | 0.32 | 0.37 |
| SOREMP (Yes/No) (MSLT), % | 22.9% | 42.6% | 30.0% | 46.6% | 0.62 | 35 | 30 | 0.60 | 0.971 |
| Sleep efficiency (PSG), % | 86.5% | 15.6% | 89.6% | 8.7% | 0.27 | 35 | 29 | 0.33 | 0.64 |
| Apnea hypopnea index (PSG), events/h | 1.56 | 1.23 | 2.05 | 2.40 | 0.41 | 36 | 29 | 0.11 | 0.68 |
| Sleep latency (PSG), min | 27.10 | 72.99 | 20.10 | 30.54 | 0.42 | 36 | 29 | 0.14 | 0.70 |
| PLMI (PSG), events/h | 0.99 | 2.51 | 1.03 | 2.12 | 0.81 | 36 | 28 | 0.86 | 0.63 |
| Arousal index (PSG), events/h | 10.16 | 3.87 | 9.84 | 3.05 | 0.84 | 35 | 29 | 0.35 | 0.39 |
| WASO (PSG), % | 9.4% | 9.9% | 6.9% | 6.5% | 0.35 | 35 | 29 | 0.95 | 0.25 |
| Total sleep time (PSG), min | 482.53 | 109.06 | 491.67 | 64.89 | 0.42 | 35 | 29 | 0.77 | 0.81 |
| Stage N1 sleep (PSG), min | 45.50 | 22.49 | 37.50 | 17.63 | 0.52 | 31 | 25 | 0.97 | 0.34 |
| Stage N2 sleep (PSG), min | 255.76 | 72.48 | 263.30 | 52.07 | 0.18 | 31 | 25 | 0.49 | 0.42 |
| Stage N3 sleep (PSG), min | 67.06 | 34.50 | 75.10 | 28.72 | 0.77 | 31 | 25 | 0.84 | 0.50 |
| REM sleep (PSG), min | 111.32 | 43.13 | 105.12 | 32.65 | 0.74 | 31 | 25 | 0.48 | 0.83 |
| Early morning N3 sleep (PSG), % | 26.7% | 8.2%* | 60.0% | 10.0%* | 0.05 | 30 | 25 | 0.067 | 0.71 |
| JESS in unmedicated conditions | 15.59 | 5.35 | 15.67 | 5.40 | 0.65 | 29 | 24 | 0.70 | 0.91 |
| MSFsc | 4:32 AM | 1 h 43 min | 4:26 AM | 1 h 56 min | 0.62 | 31 | 25 | 0.77 | 0.75 |

Data are presented as the mean and standard deviation (SD), except where indicated by an asterisk (*), which denotes standard error of the mean (SE).

MSLT: multiple sleep latency test; SOREMP: sleep onset rapid eye movement period; PSG: polysomnography; PLMI: periodic limb movement; WASO: wake after sleep onset; REM: rapid eye movement; Early morning N3 sleep: N3 within 2 h prior to the scheduled wake time; JESS: Japanese version of the Epworth Sleepiness Scale; MSFsc: midpoint of sleep-corrected for free days.

Supplementary Table S12. Results of MR analyes.

| | | | | | | | | | | IVW Cochran's Q | | | MR-Egger | |
|---|---|---|---|---|---|---|---|---|---|---|---|---|---|---|
| Exposure | Instruments for *P* value threshold | Mean *F*-statistic | Methods | Beta | SE | OR | OR_L95 | OR_U95 | *P* value | Statistic | $I^2$ | *P* | Intercept | *P* |
| UKB ukb-b-4956 | $6×10^{-9}$ | 49.88 | IVW | 0.042 | 0.013 | 1.04 | 1.02 | 1.07 | 0.0013 | 67.67 | 0.00% | 0.71 | -0.0070 | 0.79 |
| | | | MR-Egger | 0.048 | 0.028 | 1.05 | 0.99 | 1.11 | 0.089 | | | | | |
| | | | WM | 0.050 | 0.022 | 1.05 | 1.01 | 1.10 | 0.025 | | | | | |
| | | | MR-PRESSO | 0.041 | 0.012 | 1.04 | 1.02 | 1.07 | 0.00094 | | | | | |
| 23andMe | $6×10^{-9}$ | 59.03 | IVW | 0.036 | 0.017 | 1.04 | 1.00 | 1.07 | 0.037 | 33.26 | 0.00% | 0.60 | -0.031 | 0.30 |
| | | | MR-Egger | 0.062 | 0.030 | 1.06 | 1.00 | 1.13 | 0.048 | | | | | |
| | | | WM | 0.047 | 0.026 | 1.05 | 1.00 | 1.10 | 0.066 | | | | | |
| | | | MR-PRESSO | 0.035 | 0.016 | 1.04 | 1.00 | 1.07 | 0.037 | | | | | |
| UKB ukb-b-4956 | $5×10^{-8}$ | 44.83 | IVW | 0.030 | 0.012 | 1.03 | 1.01 | 1.06 | 0.010 | 105.12 | 0.11% | 0.48 | -0.028 | 0.22 |
| | | | MR-Egger | 0.060 | 0.027 | 1.06 | 1.01 | 1.12 | 0.027 | | | | | |
| | | | WM | 0.049 | 0.021 | 1.05 | 1.01 | 1.09 | 0.017 | | | | | |
| | | | MR-PRESSO | 0.030 | 0.011 | 1.03 | 1.01 | 1.05 | 0.010 | | | | | |
| 23andMe | $5×10^{-8}$ | 52.09 | IVW | 0.036 | 0.017 | 1.04 | 1.00 | 1.07 | 0.031 | 40.54 | 0.00% | 0.80 | -0.026 | 0.31 |
| | | | MR-Egger | 0.061 | 0.029 | 1.06 | 1.00 | 1.13 | 0.045 | | | | | |
| | | | WM | 0.047 | 0.026 | 1.05 | 1.00 | 1.10 | 0.067 | | | | | |
| | | | MR-PRESSO | 0.035 | 0.015 | 1.04 | 1.01 | 1.07 | 0.022 | | | | | |

SE: standard error; OR: odds ratio; OR_L95: Lower limit of the 95% confidence interval for the odds ratio; OR_U95: Upper limit of the 95% confidence interval for the odds ratio;

IVW: inverse-variance weighted; WM: weighted median method; MR-PRESSO: Mendelian randomization pleiotropy residual sum and outlier.

Supplementary Table S13. Comparisons of MR results (sensitivity analyses) for chronotype exposure and IH defined by 24-h PSG >=660 min and MSLT.

| Methods | Exposure (Instruments for *P* value threshold) | 24hPSG_IH (>=660min) | | | MSLT_IH | | | | |
|---|---|---|---|---|---|---|---|---|---|
| | | Beta | SE | *P* | Beta | SE | *P* | $Z^2$ | *P* (z-test) |
| MR-Egger | UKB ukb-b-4956 ($P<5\times10^{-8}$) | 0.103 | 0.062 | 0.102 | 0.043 | 0.029 | 0.142 | 0.477 | 0.633 |
| | 23andMe ($P<5\times10^{-8}$) | 0.101 | 0.069 | 0.149 | 0.047 | 0.032 | 0.149 | 0.424 | 0.672 |
| | UKB ukb-b-4956 ($P<6\times10^{-9}$) | 0.122 | 0.071 | 0.089 | 0.029 | 0.033 | 0.377 | 0.233 | 0.816 |
| | 23andMe ($P<6\times10^{-9}$) | 0.120 | 0.077 | 0.129 | 0.051 | 0.036 | 0.158 | 0.481 | 0.631 |
| WM | UKB ukb-b-4956 ($P<5\times10^{-8}$) | 0.066 | 0.041 | 0.111 | 0.036 | 0.024 | 0.134 | 0.297 | 0.767 |
| | 23andMe ($P<5\times10^{-8}$) | 0.114 | 0.061 | 0.061 | 0.035 | 0.028 | 0.217 | 0.381 | 0.703 |
| | UKB ukb-b-4956 ($P<6\times10^{-9}$) | 0.100 | 0.048 | 0.038 | 0.040 | 0.027 | 0.144 | 0.475 | 0.635 |
| | 23andMe ($P<6\times10^{-9}$) | 0.120 | 0.065 | 0.066 | 0.038 | 0.031 | 0.219 | 0.393 | 0.694 |
| MR-PRESSO | UKB ukb-b-4956 ($P<5\times10^{-8}$) | 0.040 | 0.022 | 0.076 | 0.024 | 0.012 | 0.052 | 0.484 | 0.628 |
| | 23andMe ($P<5\times10^{-8}$) | 0.079 | 0.035 | 0.030 | 0.021 | 0.017 | 0.211 | 0.290 | 0.772 |
| | UKB ukb-b-4956 ($P<6\times10^{-9}$) | 0.051 | 0.025 | 0.042 | 0.040 | 0.014 | 0.007 | 1.428 | 0.153 |
| | 23andMe ($P<6\times10^{-9}$) | 0.081 | 0.038 | 0.043 | 0.024 | 0.020 | 0.243 | 0.247 | 0.805 |

SE: standard error; PSG: polysomnography; MSLT: multiple sleep latency test; UKB: UK Biobank; WM: weighted median method; MR-PRESSO: Mendelian randomization pleiotropy residual sum and outlier.

Supplementary Table S14. Predictive performance of UK Biobank-based sleep trait PRSs in white British and East Asian populations.

| Trait Name | Population | Model | Predictive Performance | | | | |
|---|---|---|---|---|---|---|---|
| | | | Evaluation Metric | Value | lower bound of 95% CI | upper bound of 95% CI | P-value |
| Sleep duration | hold-out test set (white British) n = 67,425 | PRS | r2 | 0.0080 | 0.0066 | 0.0093 | 6.5E-119 |
| Sleep duration | East Asian n = 1,704 | PRS | r2 | 0.0070 | -0.0009 | 0.015 | 6.9.E-04 |
| Getting up in morning | hold-out test set (white British) n = 67,425 | PRS | r2 | 0.011 | 0.010 | 0.013 | 1.7E-165 |
| Getting up in morning | East Asian n = 1,704 | PRS | r2 | 0.010 | 0.001 | 0.019 | 4.7E-05 |
| Morning/evening person (chronotype) | hold-out test set (white British) n = 67,425 | PRS | r2 | 0.031 | 0.028 | 0.034 | 0 |
| Morning/evening person (chronotype) | East Asian n = 1,704 | PRS | r2 | 0.018 | 0.005 | 0.030 | 2.3E-07 |
| Nap during day | hold-out test set (white British) n = 67,425 | PRS | r2 | 0.017 | 0.015 | 0.019 | 1.0E-258 |
| Nap during day | East Asian n = 1,704 | PRS | r2 | 0.0095 | 0.0003 | 0.0186 | 8.1E-05 |
| Daytime dozing / sleeping | hold-out test set (white British) n = 67,425 | PRS | r2 | 0.0054 | 0.0043 | 0.0065 | 2.2E-81 |
| Daytime dozing / sleeping | East Asian n = 1,704 | PRS | r2 | 0.0024 | -0.0022 | 0.0071 | 0.046 |

Results include $r^2$ values, 95% confidence intervals (CI), p-values, and sample sizes. All PRSs were originally constructed by Tanigawa et al (https://doi.org/10.1371/journal.pgen.1010105).
